# Application of graph models to the identification of transcriptomic oncometabolic pathways in human hepatocellular carcinoma

**DOI:** 10.1101/2024.04.30.24306608

**Authors:** Sergio Barace, Eva Santamaría, Stefany Infante, Sara Arcelus, Jesús De la Fuente, Enrique Goñi, Ibon Tamayo, Idoia Ochoa, Miguel Sogbe, Bruno Sangro, Mikel Hernaez, Matías A. Ávila, Josepmaria Argemi

**Affiliations:** DNA and RNA Medicine Division, Applied Medical Research Center (CIMA), University of Navarre, 31008, Pamplona, Spain; Centro de Investigación Biomédica en Red de Enfermedades Hepáticas y Digestivas (CIBER-EHD), 28029, Madrid, Spain; Facultad de Medicina Humana, Universidad de Piura, 15074, Lima, Peru; Bioinformatics Platform, CIMA, University of Navarre, 31008, Pamplona, Spain; TECNUN, Universidad de Navarra, Pamplona, Spain; Liver Unit, CUN, University of Navarre, 31008, Pamplona, Spain; Instituto de Investigación Sanitaria de Navarra (IdisNA), Pamplona, Spain; Solid Tumor Program, Hepatology Laboratory, Applied Medical Research Center (CIMA), University of Navarre, 31008, Pamplona, Spain; Division of Gastroenterology Hepatology and Nutrition, University of Pittsburgh, Pittsburgh, PA, 15232, USA

**Keywords:** Hepatocellular carcinoma, RNA sequencing, metabolism, signature, graph, gene set enrichment analysis, gene set variation analysis

## Abstract

Whole tissue transcriptomic analyses have been helpful to characterize molecular subtypes of hepatocellular carcinoma (HCC). Metabolic subtypes of human HCC have been defined, yet whether these different metabolic classes are clinically relevant or derive in actionable cancer vulnerabilities is still an unanswered question. Publicly available gene sets or gene signatures have been used to infer functional changes through gene set enrichment methods. However, metabolism-related gene signatures are poorly coexpressed when applied to a biological context. Here, we apply a simple method to infer highly consistent signatures using graph models. Using The Cancer Genome Atlas Liver Hepatocellular cohort (LIHC), we describe the main metabolic clusters and their relationship with commonly used molecular classes, and with the presence of TP53 or CTNNB1 driver mutations. We find similar results in our validation cohort, the LIRI-JP cohort. We describe how previously described metabolic subtypes could not have therapeutic relevance due to their overall downregulation when compared to non-tumoral liver, and identify N-Glycan, Mevalonate and Sphingolipid biosynthetic pathways as the hallmark of the oncogenic shift of the use of Acetyl-coenzyme A in HCC metabolism. Finally, using DepMap data, we demonstrate metabolic vulnerabilities in HCC cell lines.

## 1. Introduction

Gene Set Enrichment (GSE) methods have been widely used to facilitate the functional interpretation of transcriptomic data using sets of selected genes that are assigned to a specific biological context (1). GSE methods such as Gene Set Variation Analysis (GSVA) have enabled the interpretation of thousands of gene expression changes between conditions or groups of patient samples by integrating statistical post hoc analysis into pathway-centric models (2). However, the functional diversity between species, organs, tissues, and cell types as well as the heterogeneity of human cohorts weakens the generalization capabilities of most published signatures and gene sets, which were likely generated in highly controlled in vitro experiments on cell types and organs not related to the conditions under investigation. Similarly, some public gene sets have been curated by experts using knowledge about a specific pathway or biological process. For instance, the Metabolic atlas (MetAtlas) repository was created from genome-scale metabolic models based on multi-omics and specific tissue subsystems (3). In the Molecular Signature Database (MSigDB) hundreds of gene set collections are obtained largely from perturbation experiments (4), limiting their use in other, less controlled, transcriptomic analyses. A generalized method to adapt the available signatures to the biological context under study is thus warranted.

Hepatocellular carcinoma (HCC) is a leading cause of cancer-related death worldwide (5,6). Hepatocyte differentiation is one of the most important prognostic factors in HCC (7), as exemplified by the histological classification first proposed by Edmondson and Steiner (8). High degree HCCs -grades II and IV of Edmonson-behave aggressively with easily distinguishable atypical cell shape. As a highly proliferative cancer the metabolism of high-histological grade HCC shifts towards a more glycolytic phenotype, with more oxidative stress and glutathione usage, and activation of the pentose phosphate pathway for the synthesis of purines and pyrimidines (9). In recent years, the availability of transcriptomic data from human HCC has allowed the application of machine learning approaches for inferring metabolic classification with prognostic value (10–13). These works have tried to understand the metabolic underpinnings of HCC in an unbiased manner, generating *de novo* signatures, mostly based only in cancer samples. Despite these analytic efforts, targeting cancer metabolic reprogramming is still an unmet objective. The performance of GSVA or other GSE methods using publicly available metabolic signatures has not been yet explored to define HCC metabolism, most probably because of the above-mentioned limitations.

In this work, to find metabolic vulnerabilities in human HCC, we developed a simple method to adapt published signatures by applying graph models to filter off-the-shelf gene sets before performing GSVA. We used the two largest available cohorts of sequenced HCC samples (TCGA-LIHC and ICGC-LIRI-JP) and showed the poor coexpression of published metabolic signatures present in MetAtlas and MSigDB.

The application of graph models led to the identification of metabolic clusters, with increased coexpression. We describe the association of newly generated metabolic signatures to other well-known transcriptomics HCC subclasses such as those of Hoshida (14), Chiang (15) and to the presence of *TP53* or *CTNNB1* driver mutations. We focus our study on signatures found to be enriched in tumors when compared to non-tumoral tissue, namely N-Glycan, Mevalonate and Sphingolipid biosynthetic pathways. Finally, we show the genetic vulnerabilities within these pathways using DepMap initiative (https://depmap.org/portal/) and suggest future avenues to target oncometabolic pathways in HCC.

## 2. Materials and Methods

### 2.1 Data collection

Raw transcriptomic counts and clinical information of TCGA-LIHC cohort were obtained from Xenabrowser platform of University of California Santa Cruz (UCSC) (https://xenabrowser.net/datapages/). Japanese raw RNA sequencing counts belonging to ICGC-LIRI-JP cohort were downloaded from ICGC Portal (https://dcc.icgc.org/projects) with its corresponding clinical information. Clinical data from the TCGA-LIHC and the ICGC-LIRI-JP cohorts are summarized in **Suppl. Table 1**. Human molecular signatures tested in this article were obtained from Metabolic Atlas repository (https://metabolicatlas.org/explore/Human-GEM/gem-browser) and Molecular Signature Database (https://www.gsea-msigdb.org/gsea/msigdb/). Cell lines used for dependency and gene effect analyses were collected from DepMap portal (https://depmap.org/portal/).

### 2.2. Normalization and filtering method

Raw counts from both cohorts were normalized using edgeR (version: 4.0.16) (16). From differential gene expression list, those genes with less expression than 1 in more than 30% of samples were removed considering as non-expressed features. Then, library size and normalization were performed by function calcNormFactors performing TMM methodology. Subsequently, counts per million (CPM) were computed from resulting normalized library sizes and used for downstream analyses.

### 2.3. Gene set adaptation based on graph models

Graph models were developed using igraph package (version: 2.0.2) (17). Using CPM from normalized cohorts and retrieved metabolic gene sets, gene-gene coexpression was calculated using Spearman correlation after noticing extreme values in some observations. Correlation matrixes were filtered removing reciprocal values contained in diagonal and selecting highly correlated genes applying different correlation cutoffs called as Correlation Cutoff of Input Matrix (CCIM). Then, filtered matrixes were used as input for graph development. In these graphs, median, variance, gene set size, centrality and Louvain communities were examined in each gene set tested. We considered eigenvalue centrality as our preferential metric for estimating the centrality of each gene in the graph in a range between 0 (isolated gene) and 1 (central gene). We further included community and membership studies to evaluate if the same gene set clustered into two or more different subsets with different biological interpretation. Therefore, we established the resolution filter from Louvain function indicating that those communities with at least 20% of the total size should be included in the output file. Additionally, for each gene set central gene and core genes with the best centrality (eigenvalue _≥_ 0.9) were collected. Successively, central gene renamed the resultant adapted gene sets compiling single or multiple gene set with the same core genes considered as a gold standard.

Next, it was indispensable to test whether adapted gene sets reflect the original biological collection. Therefore, each gene set was assessed with clusterProfiler package (version: 4.10.1) (18) performing hypergeometric test. Any adapted gene set that was first-ranked in hypergeometric test was included as it indicated that biological information was maintained, otherwise it was removed. Finally, a set of selected and enriched gene sets were collected for downstream analyses.

### 2.4. Sample enrichment with ssGSEA

Single sample Gene Set Enrichment algorithm (ssGSEA) from corto package (version: 1.2.4) (19) was performed to estimate the enrichment of each gene set across non tumor and tumor samples. Those ones with less than 2 genes where removed from ssGSEA enrichment.

### 2.5. Statistical analysis based on ssGSEA score

Molecular and metabolic HCC subgroups obtained from previous works (10,14,15) and sample information presented in clinical data were evaluated considering the score of ssGSEA algorithm. Those samples whose initial diagnosis was not related to Hepatocellular carcinoma were removed. From TCGA-LIHC, a total of 359 tumor samples and 49 non-tumor samples with transcriptomic and metadata information were used for statistical analyses. Mean, standard deviation and statistical test were performed and retrieved for posterior visualization.

### 2.6. Ridgeplot and heatmap visualization

Statistically significant gene sets found in molecular and clinical categories were plotted with pheatmap (version: 1.0.12) (20), ggplot2 (version: 3.5.0), ggridges (version: 0.5.6) and ggthemes (version: 5.1.0) (21). When analysing differences of gene set enrichments between non-tumor and tumor samples, only paired samples were selected. Hoshida, Chiang, iHCC and main mutations (TP53 and CTNNB1) where compared with non-tumor samples. Samples without information in any category were removed from the analyses.

### 2.7. Survival analysis

TCGA patients were previously divided into two partitions (training and validation subsets) with 50:50 proportion assuming the presence of overfitting phenomenon. For both subsets, survival analysis was performed taking median ssGSEA score as numerical discriminator between High and Low-expressed groups. Median survival of both groups was estimated using survival (version: 3.5-5) and survminer (version: 0.4.9) packages (22,23). In addition, Median Survival Difference (MSD) was computed subtracting Low to High-expressed samples.

### 2.8. Validation

For ICGC-LIRI-JP samples, adapted gene sets obtained from TCGA-LIHC samples were validated. Following methodology of previous sections (2.4 to 2.7), it was evaluated the similarity between TCGA and LIRI samples. Only those samples reported as Hepatocellular Carcinoma were selected for downstream analysis compelling 172 non-tumor and 200 tumor samples. ICGC-LIRI-JP was considered as test subset in survival analyses.

### 2.9. Estimation of gene dependency and gene effect with DepMap Portal

Cell lines from HCC (**Suppl. Table 2**) were in silico tested using our metabolic gene collection to estimate gene effect and gene dependency in each cell line. To this end, we downloaded the CRIPR-Cas9 screen database from DepMap portal (29083409, 31068700). Null score leads to minimal dependency effect on cell line whereas positive score indicated cell dependency. Regarding gene effect score, null score represented no cell dependency while negative scores indicated deleterious effect on the cell line.

### 2.10. Statistical analysis

Shapiro-Wilk test was used to test the normality of each distribution. For variables with two groups, T-test or Wilcoxon test was performed according to normality test. For three or more groups, parametric ANOVA or Kruskal Wallis tests were performed and each individual comparison two by two were also evaluated. Log-rank test was used for survival analyses. In boxplots, median and interquartile range were displayed. Only those p values less than 0.05 were considered statistically significant.

## 3. Results

### 3.1. Graph models generate highly compacted metabolic signatures

Considering metabolism as a crucial hallmark of HCC development, we first investigated how co-expressed curated signatures from Hallmark collection of MSigDB are in the context of the HCC transcriptome (**Fig 1A**). As expected, genes belonging to proliferative signatures such as E2F_TARGETS, describing the high protein translation rate, or MYC_TARGETS −60% of HCCs in TCGA overexpress MYC oncogene-have high median gene to gene correlation (MGGC). Metabolic signatures presented moderate to low co-expression, with MGGC values closer to non-hepatic non-cancer signatures, which presented the lowest co-expression. When the Metabolic Atlas (MetAtlas) repository was tested, similarly low levels of MGGC were found (**Fig 1B**), with higher values in signatures related to oxidative phosphorylation or beta-oxidation of fatty acids when compared to others such as acylglycerides metabolism or cytosolic carnitine shuttle. These data confirmed the specificity of the biological context of HCC and suggest that enrichment scores based on these signatures could be affected by low signal to noise ratio.

**Figure 1.**
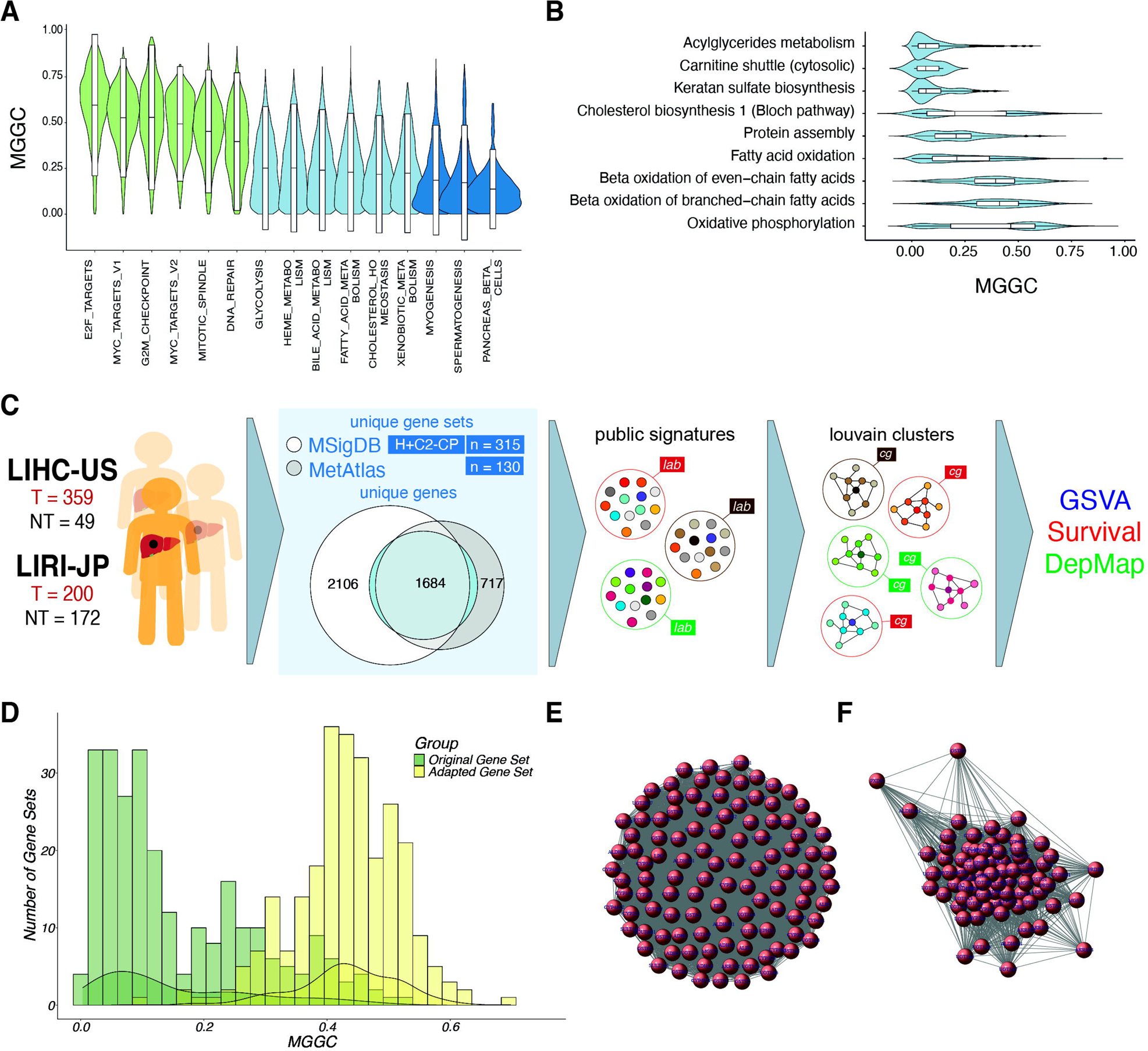
Using graph models to adapt public signatures. (A) Violin plot showing the median gene to gene correlation (MGGC) of selected Hallmark signatures in the LIHC HCC cohort, including those related with proliferation (green), metabolism (light blue) and unrelated to liver or liver cancer (dark blue). (B) MGGC of signatures from The Metabolic Atlas (MetAtlas). (C) Scheme of the method of adaptation of public signatures from Molecular Signature Database (MSigDB) and MetAtlas to identify centric nodes and metabolic clusters using graph models. (D) Effect of the method on the MGGC of metabolic signatures from MSigDB and MetAtlas in LIHC. (E) An example of a non-filtered coexpression matrix of “Xenobiotic Metabolism” Signature of the MetAtlas in the LIHC cohort, were all genes (nodes) are connected by apparently equal relationship (edge). (F) An example of a Louvain cluster obtained by after graph model was applied to “Xenobiotic Metabolism” signature.

We thus designed a pipeline to derive highly coexpressed signatures from public gene sets, using graph models, aiming at identifying networks of genes based on co-expression matrices from human HCC transcriptomic data (see Methods). To pursue this, we retrieved 130 metabolic signatures from Metabolic Atlas and 315 Hallmark (H) and canonical pathways from curated gene sets (C2-CP), from MSigDB (**Fig 1C**). A total of 445 metabolic signatures were analyzed with 1684 genes shared in both databases, 2105 uniquely present in MetAtlas and 717 specific of the MSigDB. As expected, globally, adapted gene sets obtained from bioinformatic pipeline presented better MGGC compared to their original counterparts (**Fig 1D**).

Next, TCGA-LIHC samples were randomly clustered into training and validation sets, while ICGC-LIRI-JP composed the test set. Graphs generated with higher correlation thresholds generated less populated graphs and smaller Louvain communities (**Fig 1E-F**). Testing all possible correlation thresholds, we detected that a range between 0.2 and 0.5 generated the highest number of communities with the highest number of coexpressed genes, with a considerable increase in MGGC and decrease in median gene-gene variance (MGGV), at expenses of a limited reduction of the gene set size to between 40 and 30% of its original size, indicating that a significant number of genes in the original public signature in MetAtlas or MSigDB was preserved (**Suppl. Fig 1A-D**). We also confirmed that all newly generated signatures were ranked first after a hypergeometric test was done against the universe of all available gene signatures (all GSEA H and C2 collections plus MetAtlas signatures). The final collection of gene signatures was named after its central gene (eigenvalue of 1 or the highest eigenvalue in the case of one original signature generating two or more new Louvain communities, see Methods). Finally, those signatures with the same central gene, some of them from different signatures representing similar metabolic pathways were merged.

### 3.2. Metabolic clusters are tumor-specific and associated with molecular subtypes in the TCGA LIHC cohort

The intermediate metabolism is one of the most important functions of the normal hepatocytes. We hypothesized that tumor communities could not be entirely coincident with non-tumor ones, regarding both the number and size of the gene sets and the specific central genes. It was found that non-tumor communities presented higher number of clusters, unique genes per cluster, unique core genes and unique central genes when compared to tumor ones (**Fig 2A-B**), and that the central genes defining clusters were mostly divergent (**Fig 2C-F**). After applying gene set adaptation and selecting enriched gene sets, 74 different signatures were obtained conforming different metabolic signatures, of which only 17 were found also in non-tumor samples (**Fig. 2F, Suppl Table 3**).

**Figure 2.**
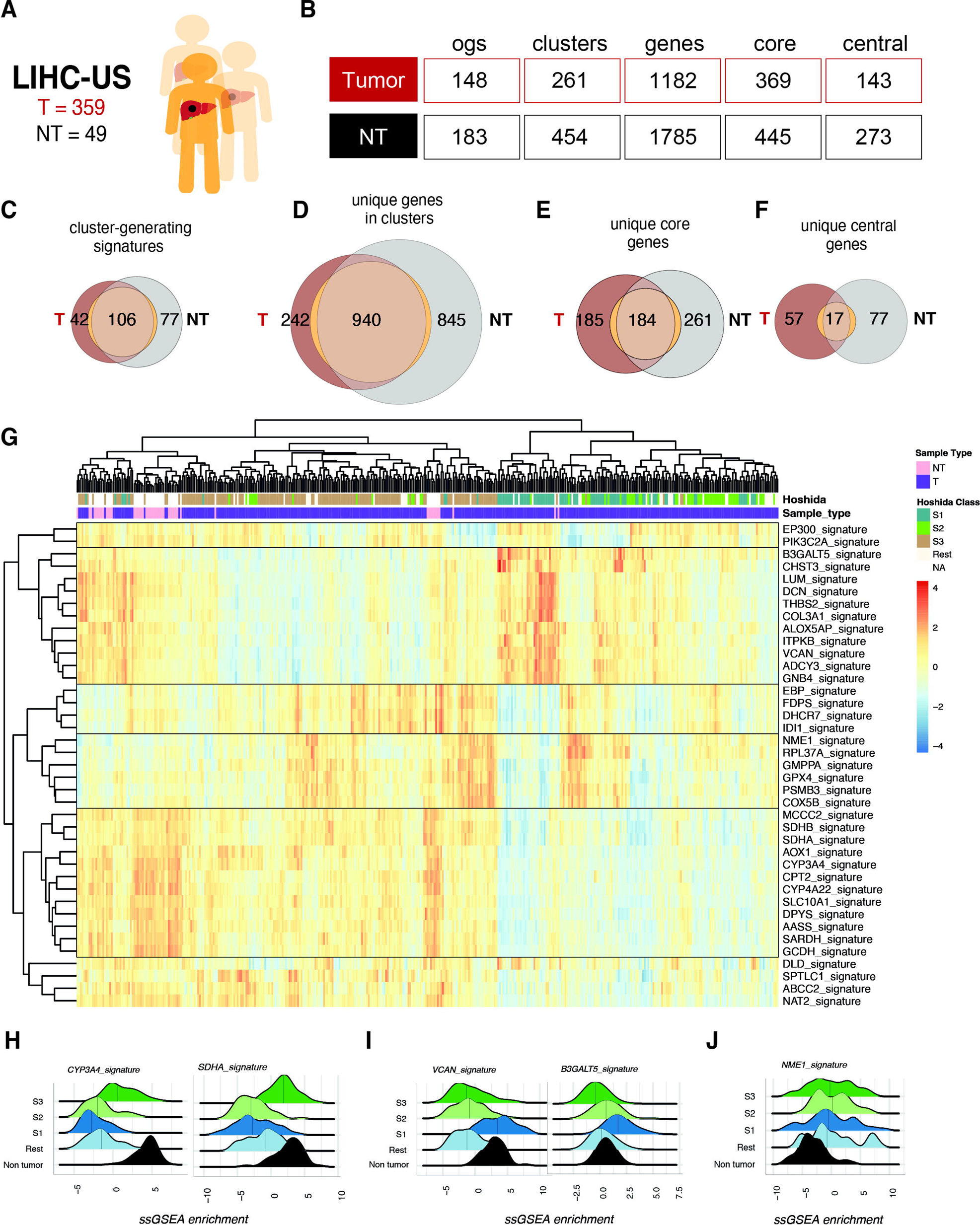
Identification of Metabolic Clusters in HCC and their association with transcriptomic classes. (A) LIHC was used as the training cohort, where tumor (HCC, n = 359) and non-tumor (NT, n= 49) samples were analyzed (B) From the original MSigDB and MetAtlas signatures, unrestricted coexpression matrix (r threshold 0.05), led to the identification of only 148 metabolic clusters in HCC and 183 in NT, which increased to 261 and 454 in HCC and NT respectively, with r threshold 0.4, used in downstream analyses. These clusters included 1182 and 1785 unique genes, 369 and 445 core genes and 143 and 273 central genes in HCC and NT respectively (see method section). (C-F) Overlap between signatures (C), unique genes (D), unique core genes (E) and unique central genes (F) found in HCC and NT. (G) Heatmap of ssGSEA scores using newly identified metabolic clusters and its association with Hoshida classes S1, S2 and S3. (H-J) Ridge plots showing the expression of signatures belonging to group 1, 2 and 3 by Hoshida Class S1, S2 and S3.

**Table 1.** Main metabolic signatures upon hierarchical clustering of TCGA-LIHC cohort.

| Hierarchical cluster | Signature name | Function |
| --- | --- | --- |
| Group 1<br>Liver-specific | CYP4A22, CPT2 | Fatty acid metabolism and transport |
|  | GCDH | Lysine, hydroxylysine and tryptophan metabolism |
|  | SARDH | Glycine cleavage |
|  | AASS | Lysine catabolism |
|  | MCCC2 | Leucine catabolism |
|  | SDHA, SDHB<br>CYP3A4, AOX1 | Respiratory chain reaction<br>Detoxification and metabolism of xenobiotics |
| Group 2<br>ECM metabolism | LUM, DCN, VCAN, THBS2, COL3A1<br>ALOXAP5<br>CHST3, B3GALT5 | Extracellular matrix, organization and cell adhesion<br>Leukotriene metabolism<br>Modification of glucosamine glycans |
| Group 3<br>Proliferation | NME1<br>RPL37A<br>COX5B<br>GPX4<br>GMPPA | Nucleotide metabolism<br>Protein synthesis<br>Oxidative phosphorylation<br>Glutathione management<br>Cytoplasmic glycosylation |
| Group 4<br>Cholesterol | IDI1, FDPS, DHCR7, EBP | Mevalonate and cholesterol biosynthesis |
| Group 5 | EP300<br>PIK3C2A | Nuclear factors<br>Inositol phosphate metabolism |
| Unclustered | DLD<br>SPTLC1<br>ABCC2<br>NAT2 | Glycolysis<br>Sphingolipid metabolism<br>Glucuronidation and transport of bilirubin<br>Metabolism of drugs |

We then decided to explore whether the relative increase or decrease of the global expression of a specific community defined a particular biology or associated with a previously described HCC subtype. We thus interrogated the enrichment of the new graph-identified signatures using ssGSEA in the HCC samples of the TCGA-LIHC cohort (**Fig 2G-K**). The hierarchical clustering of the signatures led to four main groups of pathways (**Fig2G, Table1**), which are described in what follows.

- The largest group of pathways included a varied array of typically hepatic metabolic functions: some related to fatty acid metabolism and transport, such as Cytochrome P450 Family 4 Subfamily A Member 22 (CYP4A22) or Carnitine Palmitoyltransferase 2 (CPT2) involved in mitochondrial long-chain fatty acids transport. Other related to the catabolism of aminoacids, such as Glutaryl-CoA Dehydrogenase (GCDH), an important enzyme in the degradation of lysine, hydroxylysine, and tryptophan, Sarcosine Dehydrogenase (SARDH) involved in glycine cleavage, Alpha-Aminoadipic Semialdehyde Synthase (AASS), in charge of lysine degradation, and the Methylcrotonoyl-CoA Carboxylase 2 (MCCC2), involved in the catabolism of leucine. Some additional pathways related to this group included those centered in enzymes of the respiratory chain, such as the subunits of the Succinate Dehydrogenase (SDHA and B), and enzymes and transporters involved in the processing of drugs and xenobiotics (CYP3A4, AOX1, NAT2 and ABCC2).
- The second largest group of pathways included functions related to metabolic aspects of extracellular matrix (ECM) organization and cell adhesion, such as signatures centered in Lumican (LUM), Decorin (DCN), Versican (VCAN), thrombospondin 2 (THBS2) and Collagen Type III Alpha 1 Chain (COL3A1); pathways related to inflammation including the leukotriene biosynthesis pathway centered in Arachidonate 5-Lipoxygenase Activating Protein (ALOXAP5); and pathways related to the modification of glucosamine glycans such as those centered in the Carbohydrate Sulfotransferase 3 (CHST3) and the Beta-1,3-Galactosyltransferase 5 (B3GALT5) genes.
- A third group was composed of signatures related to nucleotide synthesis such as Nucleoside Diphosphate Kinase 1 (NME1), protein synthesis (RPL17A-driven signature), mitochondrial function (COX5B), glutathione (GPX4) and cytoplasmic glycosylation pathways (GMPPA).
- The fourth group included gene sets related to mevalonate and cholesterol biosynthesis, such as Isopentenyl-diphosphate Delta Isomerase 1 (IDI1), Farnesyl Diphosphate Synthase (FDPS), 7-Dehydrocholesterol Reductase (DHCR7) and the Emopamil-Binding Protein (EBP).
- Interestingly, a residual group with ssGSEA values unrelated to any of the four mentioned groups encompassed transcriptional regulators and nuclear factors such as the ones included in the mediator complex and nuclear correpressors included in the EP300 community, and a PIK3C2A-centered signature, with genes involved in the inositol-phosphate metabolism.

Interestingly, some of the Hoshida subtypes, originally not defined based on the metabolic characteristics (14), clustered according to some of the new graph-based metabolic signatures (**Fig 2G**, top). For example, while Hoshida S1 class was enriched in group 2 signatures (Metabolism of ECM related proteins), S3 class had higher scores of group 1 (Liver-specific). Both signatures are present in the non-tumoral tissue, which suggests that in S1 tumors, there is a specific downregulation of group 1 signatures, such as CYP3A4 and SDHA (**Fig 2H**), and in S3 tumors there is a downregulation of group 2, such as VCAN or B3GALT5 (**Fig 2I**). As expected, the intensity of liver-specific signatures was higher in non-tumoral liver than in S3 tumors (**Fig2H**). On the other hand, the intensity of ECM signatures was slightly higher in S1 tumors than in non-tumors (**Fig2I**). Hoshida class S2 has lower expression of both group 1 and group 2 signatures when compared to non-tumor samples (**Fig 2G-I**). Whether group 1 and group 2 signatures reflect two differentiation states is unknown. Two groups of metabolic signatures (group 3, “proliferation” and group 4, “cholesterol”) had increased ssGSEA scores in tumor samples when compared to non-tumoral livers, indicating potential metabolic targets. Samples with high scores in group 3 and 4 signatures were distributed among Hoshida S1, S2 and S3 classes. This could indicate that while liver differentiation and ECM define one layer of metabolic subtype, the proliferation rate – in terms of protein synthesis and nucleotide and mitochondrial metabolism- and the induction of the mevalonate pathway could define a second perhaps more dynamic onco-metabolic shape (**Fig 2J-K**). Interestingly, not all tumors with high group 3 had increased group 4 signature scores, indicating different regulatory control.

We then analyzed Chiang transcriptomic classes (15) in the context of the newly generated metabolic signatures. As with Hoshida subclasses, there were metabolic differences between Chiang subclasses. For example, proliferative subgroup presented the lowest levels of expression of group 1 signatures, whereas CTNNB1, Polysomy 7 and Interferon classes presented higher expression of these signatures (**Suppl. Fig. 2A**). Conversely, when interrogating group 2 signatures, CTNNB1 and Polysomy 7 seven, but not the Interferon class had the lowest level of expression (**Suppl. Fig. 2B**). These results indicated the relationship between a specific molecular subtype and the metabolic status of the cell.

Finally, the metabolic landscape of HCC as inferred by the generated signatures matched with previously described metabolic classes. Bidkhori metabolic class iHCC1 (10) was enriched with liver-specific (group 1) signatures (**Suppl. Fig. 2C and D**), while iHCC3 tumors had the lowest levels of these signatures but highest of metabolism of ECM (**Suppl. Fig. 2C and E**). As described, iHCC2 metabolic subgroup had an intermediate score in group 1 signatures when compared to iHCC1 and 3. Some signatures such as those related to drug and xenobiotic metabolism (NAT2, UGT1A4) and those related to steroid metabolism (HSD17B4) (**Suppl. Fig. 2C and F**).

### 3.3. TP53 and CTNNB1 mutant tumors are metabolically diverse

HCC main driver mutations include deleterious *TP53* variants, activating N terminal *CTNNB1* mutations and activating variants of *TERT* gene promoter. Among them, *TP53* and *CTNNB1* mutations are mutually exclusive in most HCC patients, which allow for the comparative analysis of their specific biologic behavior. With newly adapted signatures, we could define a specific metabolic shape for *TP53* and *CTNNB1* mutated tumors. Tumors bearing *TP53*-null mutations presented overexpression of NME1 signature, related with purine metabolism. On the other hand, *CTNNB1*-mutated patients presented an enrichment in metabolic signatures such as *MAT1A* and *CYP3A4*, when compared to *TP53*-null. Conversely tumors with wild type *TP53* presented higher metabolic enrichment of *MAT1A* and *CYP3A4*. These data suggest that *CTNNB1* mutation supports the maintenance of a liver metabolic-like phenotype in HCC, while *TP53* mutant tumors are more de-differentiated and highly proliferative (**Suppl. Fig. 3**).

### 3.4. The survival of patients with low metabolic tumors is worse in the LIHC and LIRI cohorts

It was previously observed that Bidkhori iHCC1 class determined survival prognosis (10). We therefore used the training, validation, and test cohorts to verify the prognostic significance of our derived metabolic signatures (**Fig 3A**). Only liver-specific metabolic signatures (group 1) such as ABAT, DMGDH and GLYAT, which were downregulated in tumors (**Fig 3B**) were associated with prognosis in all three cohorts. Patients with tumors having higher expression of these metabolic signatures, had increased overall survival (**Fig 3C**). This result indicates the validity of the signatures found in LIHC cohort to interrogate the metabolic phenotype of unseen data, such as the LIRI-JP cohort. The median survival difference was lower in the LIRI-JP, perhaps due to the better global survival in the cohort.

**Figure 3.**
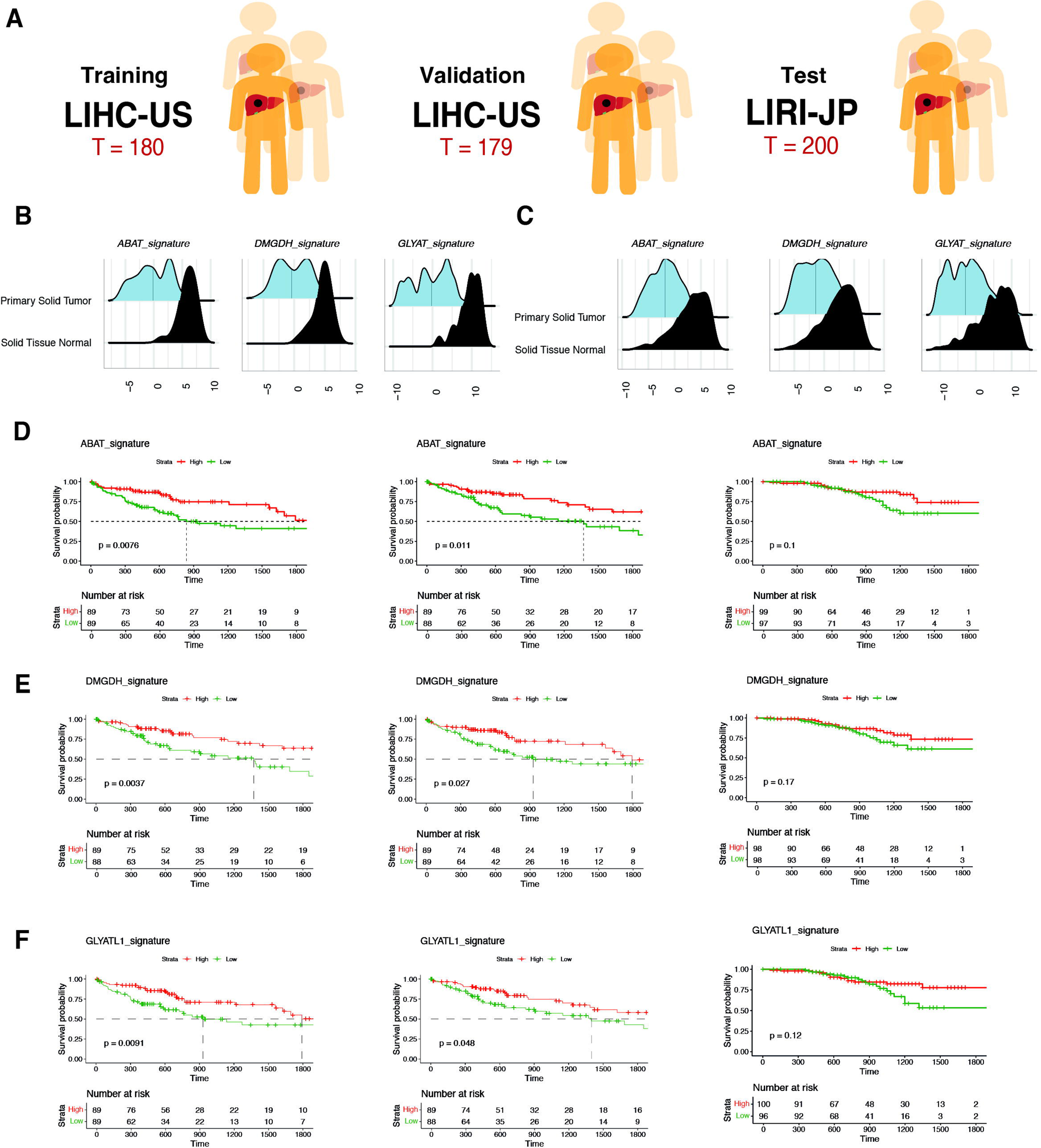
Validation of the method for prognostic prediction in patients with HCC. (A) Random 50:50 split of the LIHC led to the training (n=180) and validation (n=179) cohorts for prognostic analyses, while LIRI-JP cohort (n=200) was used as test cohort. (B-C) Overall ssGSEA scores of prognostic signatures ABAT, DMGDH and GLYAT when comparing tumor vs non-tumor in LIHC (C) and LIRI-JP (D). (E-F) Survival analyses of patients in LIHC-training, LIHC-validation and LIRI-JP-testing cohorts when dividing the population in high and low ssGSEA scores for ABAT (E), DMGDH (F) and GLYAT (G) signatures.

### 3.5. Mevalonate, N-Glycan and Sphingolipid biosynthesis pathways shape the tumor metabolism in human HCC

We then decided to study in more detail the few signatures with increased scores in tumors when compared to non-tumoral livers in both cohorts LIHC and LIRI, regardless their molecular subtype (signature groups 3 and 4). These signatures were related to the mevalonate/cholesterol biosynthetic pathway such as IDI1 signature, glycosylation (GMPPA signature), sphingolipid (SPTLC1) and nucleotide (NME1 signature) metabolism and the catabolism of the polyamines, with mostly proteasome subunit genes (PSMB3 signature). Isopentenyl-diphosphate delta-isomerase 1 (IDI1) was the most centric gene of a Louvain community with increased enrichment scores (**Fig. 4A**) involving other cholesterol-related genes such as Squalene Epoxidase (SQLE), Mevalonate Diphosphate Decarboxylase (MVD) or Sterol Regulatory Element Binding Transcription Factor 2 (SREBP2) and Farnesyl Diphosphate Synthase (FDPS), Phosphomevalonate Kinase (PMVK), all of them with increased expression in HCC of both the LIHC and the LIRI-JP cohorts (**Fig. 4B-C**).

**Figure 4.**
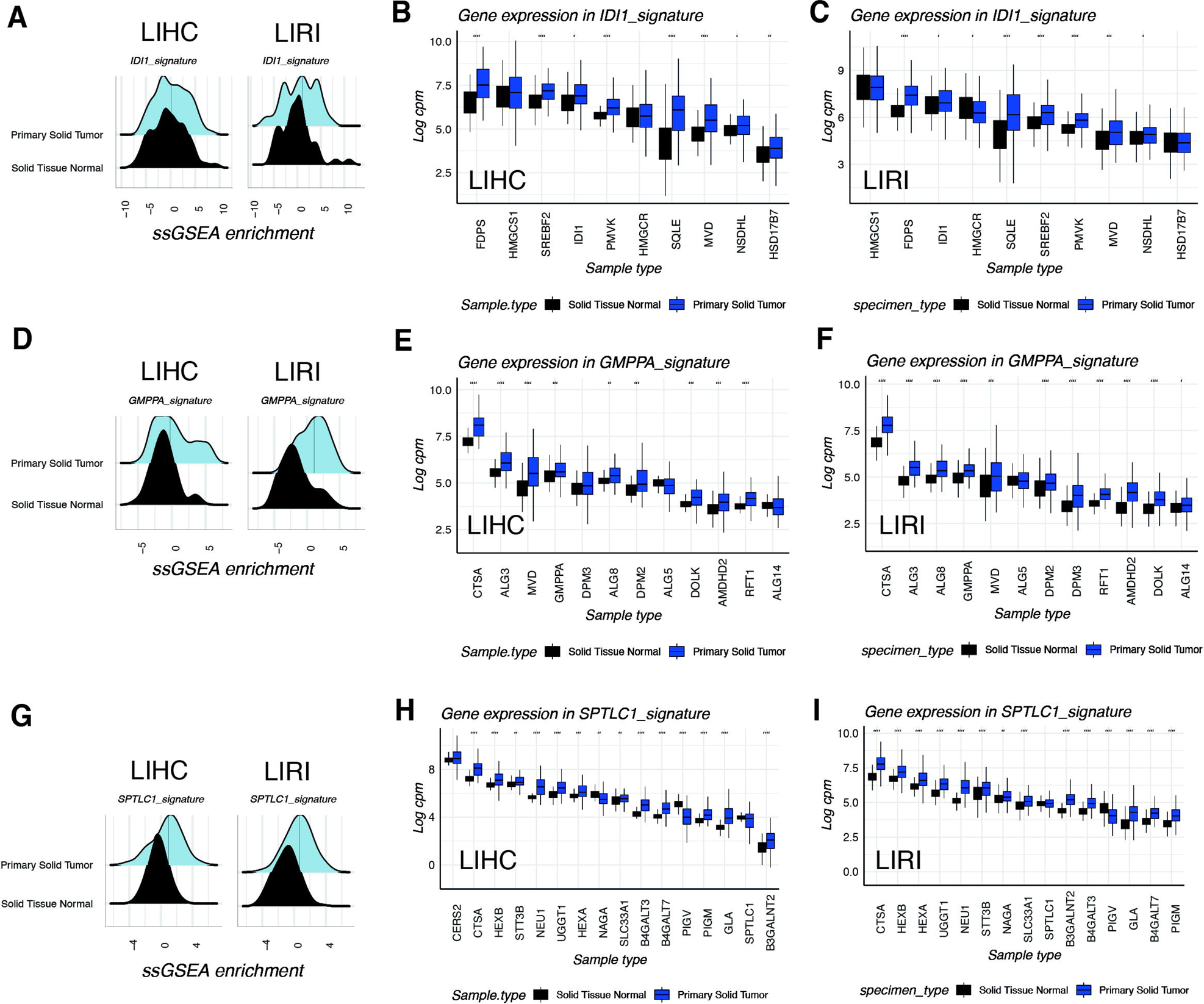
IDI1, GMPPA and SPTLC1-centered clusters are overexpressed metabolic signatures in HCC. (A) Ridge plots of ssGSEA scores of Isopentenyl-diphosphate delta-isomerase 1 (IDI1) in LIHC and LIRI-JP cohorts comparing it with paired non-tumor tissue (B-C) Box plots showing the expression levels of individual genes included in IDI1 signature in LIHC (B) and LIRI-JP (C) cohorts. (D) Ridge plots of ssGSEA scores of GDP-mannose pyrophosphorylase A (GMPPA) in LIHC and LIRI-JP cohorts comparing it with paired non-tumor tissue (E-F) Box plots showing the expression levels of individual genes included in GMPPA signature in LIHC (E) and LIRI-JP (F) cohorts. (G) Ridge plots of ssGSEA scores of Serine Palmitoyltransferase Long Chain Base Subunit 1 (SPTLC1) in LIHC and LIRI-JP cohorts comparing it with paired non-tumor tissue (H-I) Box plots showing the expression levels of individual genes included in GMPPA signature in LIHC (H) and LIRI-JP (I) cohorts.

GDP-mannose pyrophosphorylase A, encoded by GMPPA gene, was the central gene in a signature associated with other enzymes related to the synthesis and ramification of N-Glycans in the cytoplasmic and luminal domains of the Endoplasmic Reticulum wall, including Dolichyl-Phosphate Mannose Synthase Subunit 3 and 8 (ALG3, ALG8), Dolichol kinase (DOLK) and the Required for FTase Activity Protein 1 (RTF1), the flippase that internalizes the glycans to be incorporated to nascent polypeptides inside the ER, all of these genes were upregulated in tumors both in LIHC and LIRI (**Fig. 4D-F**).

The signature centered in the Serine Palmitoyltransferase Long Chain Base Subunit 1 (SPTLC1) also is upregulated in tumors of both TCGA-LIHC and ICGC-LIRI-JP cohorts (**Fig. 4G**). SPTLC1 is a key enzyme in sphingolipid biosynthesis, catalyzing the generation of ketosphingoids from Serine and Acetyl-CoA, the rate limiting step for the generation of ceramides. Other enzymes in the same signature belonged to the lysosomal pathway of ganglioside catabolism, such as Hexosaminidase Subunit Beta (HEXB) and Neuraminidase 1 (NEU1) were also overexpressed in tumor samples of both LIHC and LIRI-JP (**Fig. 4H-I**).

NME1 signature comprised nucleotide metabolism and polimerase enzymes related to DNA replication and transcription. As expected, tumor samples presented higher expression in comparison with non-tumor ones (**Suppl. Fig. 4A**). Among the most relevant genes in this signature were Thymidine Kinase 1 (TK1), Uridine-Cytidine Kinase 2 (UCK2) and Deoxythymidylate Kinase (DTYMK), all involved in nucleotide metabolism (**Suppl. Fig. 4B-C**).

Finally, overexpression of the PSMB3 signature, related to the metabolism of polyamines but also to the more general proteasome function, was found differentially increase tumor samples (**Suppl. Fig. 5A**). Several components and subunits of the proteasome presented a general upregulation in tumor samples of both cohorts LIHC and LIRI-JP (**Suppl. Fig. 5B-C**).

### 3.6. HCC metabolic vulnerabilities in mevalonate, N-glycan and sphingolipid pathays as new targets for therapy

The aforementioned results pointed to a possible implication of mevalonate/cholesterol, N-glycan and sphingolipid metabolism in HCC biology, regardless of the molecular subtype or the driver mutation. Since these pathways are induced in tumor samples when compared to non-tumoral liver, we though they could constitute potential targets for anticancer therapy. Thus, to determine the importance of these enzymes for the survival of HCC cells, DepMap data was used to analyse cell viability and dependency when these genes are targeted. After analysing 6 tumor-specific signatures (IDI1, GMPPA, NME1, PSMB3, SPTLC1 and EBP), the gene effect and gene dependency were measured in different non-cancerous cell lines and liver cancer lines of human HCC. The genes 3-Hydroxy-3-Methylglutaryl-CoA Synthase 1 (HMGCS1), 3-Hydroxy-3-Methylglutaryl-CoA Reductase (HMGCR), Farnesyl Diphosphate Synthase (FDPS) and Mevalonate Diphosphate Decarboxylase (MVD) were the most affected genes upon CRISPR-knock out, impacting on cells survival as exemplified by the highest gene dependency scores (**Fig. 5A**) and the most negative gene effect (**Fig. 5B**). Interestingly the knock down of another gene in the same community, the NAD(P)H Steroid Dehydrogenase-Like (NSDHL) conferred higher survival and have a positive effect in cancer cells (**Fig. 5B**).

**Figure 5.**
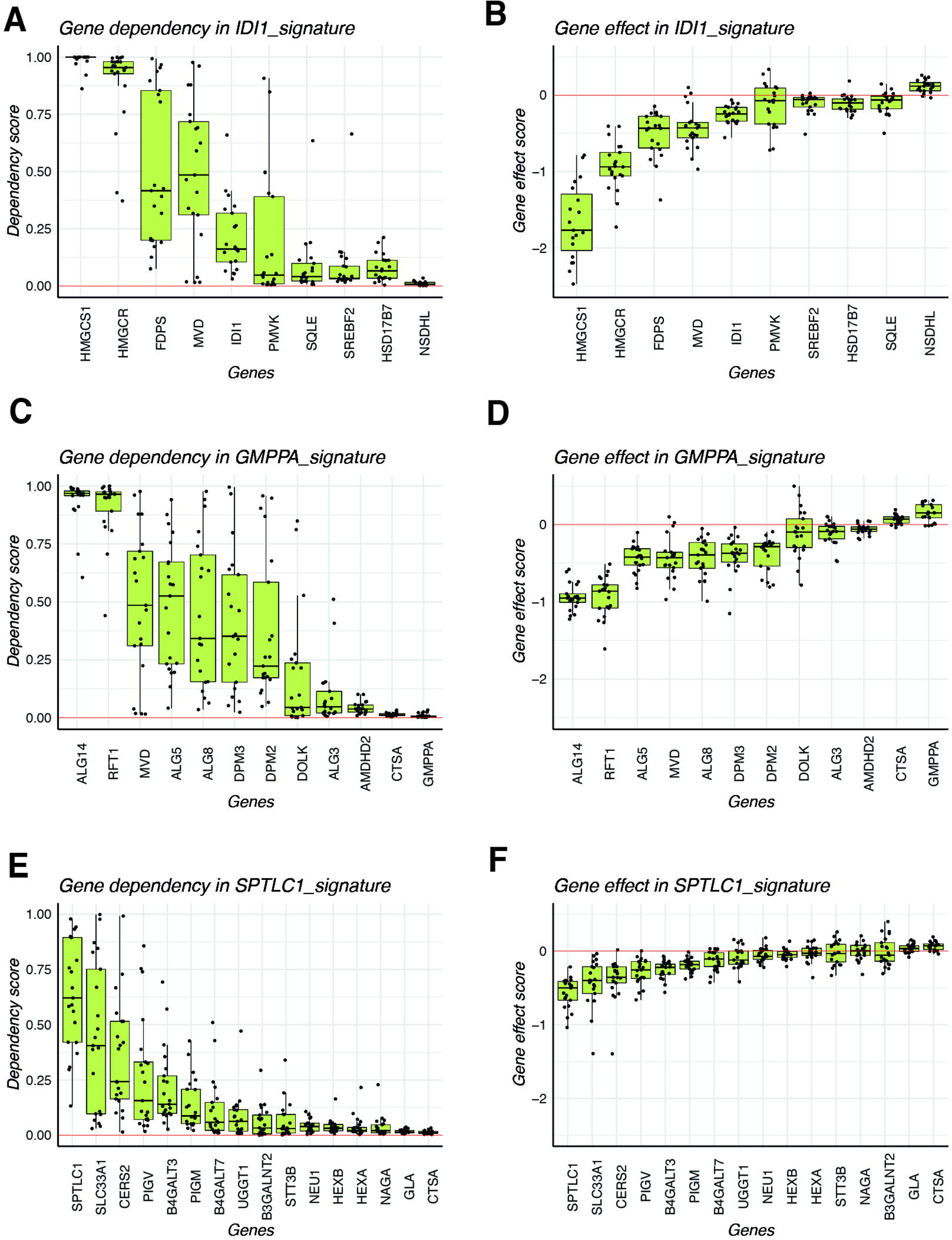
Identification of metabolic vulnerabilities in HCC cell lines using DepMap. (A) Gene Dependency scores for individual genes included in IDI1 signature. (B) Gene Effect scores for individual genes included in IDI1 signature. (C) Gene Dependency scores for individual genes included in GMPPA signature. (D) Gene Effect scores for individual genes included in GMPPA signature. (E) Gene Dependency scores for individual genes included in SPTLC1 signature. (F) Gene Effect scores for individual genes included in SPTLC1 signature.

In the N-Glycan GMPPA signature, UDP-N-Acetylglucosaminyltransferase Subunit (ALG14) and RFT1 Gene (RFT1) accounted for the highest vulnerability in most HCC cell lines tested (**Fig. 5 C-D**). In the case of SPTLC1 signature, some but not all HCC cell lines were dependent on SPTLC1 and SLC33A1 genes, indicating a pathway less relevant for HCC survival than the previous two.

## 4. Discussion

In the present work we implement a simple computational method for inferring metabolic pathways, using public signatures in MSigDB and the MetAtlas, by understanding HCC-specific gene networks using graph-models. The newly generated gene communities are highly coexpressed and represent major metabolic domains of liver cancer cells. We use ssGSEA as an enrichment method to infer the activity of these pathways in samples of the two largest cohorts with available transcriptomic data, the TCGA-LIHC and the ICGC-LIRI-JP, including a total of 559 HCC samples and 149 non-tumoral liver samples.

We show that the metabolic phenotype is only partly associated with previously described signatures such as Hoshida and Chiang. As expected, liver-enriched metabolic pathways were associated with Hoshida S3 subclass and Chiang subclasses CTNNB1, Poly7, and Interferon. On the other hand, ECM metabolism was associated with Hoshida S1. We nevertheless describe the presence of two groups of signatures, groups 3 “proliferation” and 4 “Mevalonate/cholesterol”, which can be increased or decreased in Hoshida subclasses S1, S2 and S3, indicating a level of metabolic regulation that works in asynchrony in regards to hepatocyte differentiation. In conclusion our work confirms and enriches previous transcriptomic classification of HCC, adding an important validation of the main findings in the LIRI-JP cohort which so far has not been used to study metabolic profiles. We show that *TP53* null and *CTNBB1* mutated tumors have divergent metabolic profiles, which is consonant with what has been previously described (24–26).

The strength of this study is our focus on signatures enriched in tumoral samples when compared to non-tumor tissues. Previous works (10,14,15) depicted the transcriptomic landscape of HCC, but did not consider the expression of metabolic signatures in non-tumoral livers. Here we show that those tumors with high liver-specific signature enrichment are still poorly differentiated when compared to non-tumoral livers. This approach helps us discover metabolic pathways increased in tumors that constitute part of the hallmarks of liver cancer and that could be targeted by future synergistic approaches with immunotherapy.

We confirm that metabolic pathways related to nucleotide biosynthesis such as NME1 signature, are related to highly proliferative tumors and have a role in HCC progression (27). We describe the mevalonate, the N-glycan and the sphingolipid biosynthetic pathways, as induced pathways in HCC and thus potential targets for therapy. Although these new findings validate previous evidence of the role of these pathways in cancer cell survival and immune evasion (35, 37–39, 45), such data is so far scarce and fragmented in HCC literature.

Regarding mevalonate biosynthesis, it has been shown that IDI1 promotes tumorgrowth (28). Interestingly, IDI1 represses CCL5 and CXCL10 expressing cells in the tumor microenvironment cells, increasing the capacity for immune evasion. On the other hand, EBP inhibitors have been shown to impair prostate cancer proliferation (29). FDPS has been largely studied in other cancers. For instance, is known its role in promoting glioblastoma growth, by recruiting tumor-associated macrophages through an increased expression of CCL20 (30). This same immunosuppressive mechanism set up by cancer cells has been demonstrated in in mouse models of beta-catenin induced HCC (31). In osteosarcoma cell lines and HeLa cells, FDPS was also able to change the ECM organization and promote proliferation and DNA repair (32). Finally, FDPS has been proposed as a biomarker of breast cancer development (33). Squalene epoxidase (SQLE) is capable to promote tumor growth by inhibiting apoptosis (34) and is able to interact with TGFb-SMAD axis to promote EMT and metastatic capacity (35). We observed, using the DepMap data, that HMGCR, HMGCS1, FDPS, MVD and IDI1 confer different degrees of vulnerability when knocked out with CRISPR-Cas9 in several human HCC cell lines. Whether available HMGCR inhibitors such as statins could be used as repurposed drugs for combination with immunotherapy remains a provocative possibility.

Regarding glycosylation pathways, still not much has been described in HCC. An abnormal glycosylation of the ectonucleotidase CD73 was found in HCC samples (36) but not in adjacent livers. More broadly, glycosylation patterns are known to be present in a variety of cancer type and contribute to its fitness and evasion from the immune surveillance (37–39). Targeting ALG14 or RFT1 led to HCC cell death in a consistent fashion expanding a variety of cancer cell lines. To determine whether targeting these genes or other members of this pathway is feasible and non-toxic for non-tumoral cells, requires further preclinical work.

Finally, for the sphingolipid biosynthesis pathway conflicting data has been reported. On one hand, SPTLC1 gene has been found to be anti-oncogenic. In colorectal cancer, the low expression of SPTLC1 leads to worse prognosis (40), in renal cell carcinoma, it inhibits cell proliferation (41) and in lymphoma patients, a mutation of SPTLC1 increasing its enzymatic activity, sensitized BCR-ABL tumors to imatinib (42). On the other hand, serum ceramides and sphingolipids such as S1P and SA1P are increased in patients with HCC but not in cirrhotic controls (43), and the blockade of sphingolipids in Huh7 and HepG2 cell lines led to increased susceptibility to sorafenib (44). More broadly, it has been described that sphingolipids are produced in higher amounts in cancer cells and that sphingosine-1-phosphate (S1P) intermediate promotes proliferation, migration and EMT (45) and regulates the interphase with other cells through the inhibition of sindecan 1 (46).

One potential unifying model to explain the relationship between the above-mentioned metabolic pathways in HCC could be the cancer-specific change in the use of Acetyl-CoA, the most used substrate of the cell for anabolic and energetic functions (**Figure 6**). In non-cancerous liver cells, Acetyl-CoA is a central metabolic intermediate and the maintenance of the Acetyl CoA pool is essential for growth, proliferation, and protein modification. Cancer cells have developed the capacity to capture acetate as an alternative source to glucose from the circulation and even from the intestinal microbiome (47,48). In the present work, we show that in human HCC many of the metabolic pathways using Acetyl-CoA, such as lipid biosynthesis, are downregulated when compared with not-tumoral tissues. This is particularly evident in highly de-differentiated tumors, while the unique genuinely overexpressed metabolic signatures are the mevalonate/cholesterol, N-glycan and sphingolipid pathways, all three meant to deviate Acetyl-CoA precursors into pro-tumoral biosynthetic pathways related to protein glycosylation, turnover and ECM organization. The fact that the main transporter of Acetyl-CoA into the lumen of ER and lysosomes, SLC33A1, is one of the most overexpressed genes in HCC, may foster its experimental evaluation as a potential target for HCC therapy.

**Figure 6.**
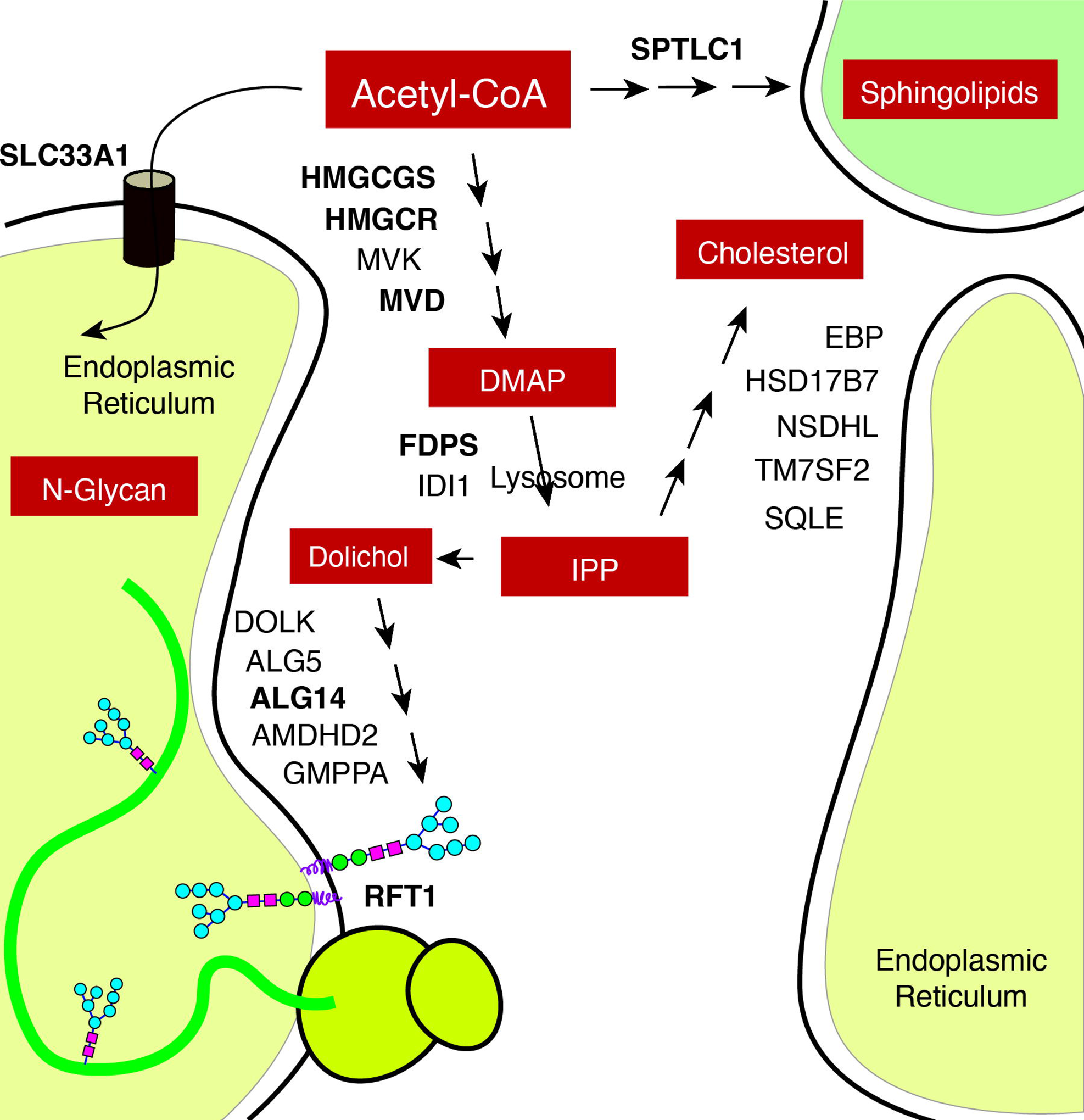
Theoretical model depicting the N-Glycan, Mevalonate and Sphingolipid biosynthetic pathways as the integrated oncometabolic responses led to the shift on Acetyl coA use in HCC.

Patients with advanced HCC are currently treated with immunotherapy in first line and two combinations are currently approved in western countries and in Asia for this indication, namely Atezolizumab plus bevacizumab or Tremelimumab plus durvalumab (REFS). One of the main limitations of this work is the lack of baseline liver and tumor transcriptomic data from patients with advanced HCC treated with these regimens. Both TCGA-LIHC and ICGC-LIRI-JP cohorts include mostly patients with early HCC that were treated through curative therapies such as resection. Whether the inhibition of the metabolic pathways found in the present work could impact in the response to these patients to immunotherapy is an area for further investigation. Also, the role of these signatures as prognostic or predictive biomarkers is yet to be explored.

## 5. Conclusions

Utilizing computational methods to infer gene networks specific to HCC, this study identifies major metabolic domains in liver cancer cells, validating and enriching previous transcriptomic classifications. The metabolic phenotype of HCC reveals associations with known HCC subclasses, such as Hoshida and Chiang, while also highlighting the presence of distinct metabolic pathways in poorly differentiated tumors compared to non-tumoral liver tissue. Mevalonate/cholesterol, N-glycan, and sphingolipid pathways emerge as potential therapeutic targets for HCC, with specific genes within these pathways showing promise for targeted therapies, potentially in combination with immunotherapy. The study also provides insights into the functional roles of various metabolic pathways in HCC progression, including nucleotide biosynthesis, glycosylation, and sphingolipid biosynthesis, shedding light on their reported involvement in tumor growth, immune evasion, and metastasis. The findings raise questions about the impact of targeting these metabolic pathways treating advanced HCC patients and suggest further investigation into the potential use of these metabolic signatures as prognostic or predictive biomarkers. Additionally, this study underscores the need for transcriptomic data from patients undergoing immunotherapy for a more comprehensive understanding of treatment responses when using current standard of care.

## Supplementary Materials

**Figure S1.** Defining the threshold for the generation of graphs; **Figure S2**. Graph-identified clusters association with iHCC metabolic classes; **Figure S3.** Selected metabolic signatures according to driver mutation (TP53 or CTNNB1); Figure S4 NME1 signature is upregulated in HCC; **Table S1**. Clinical variables of TCGA-LIHC and ICGC-LIRI-JP cohorts; **Table S2.** Characteristics of HCC cell lines from DepMap Portal; **Table S3.** Table summary of gene set adaptation in published metabolic signatures.

## Author Contributions

J.A. and M.H. conceived the idea and planned the work. J.A., B.S., and M.A.A monitored the progress of the work. S.B. did most of the in-silico work with the essential contribution by J. F., E.G. and I.T. The draft of the manuscript was reviewed by E.S., S.A., S.I., M.S., I.O, and C.A, which made critical comments and contributions. J.A., M.H. and S.B. wrote the manuscript and designed the figures. J.A., M.A.A., and M.H. share senior authorship.

## Data Availability Statement

The scripts to adapt public signatures to an expression matrix using graph models are available in GitHub (https://github.com/unav-hcclab/gsadapt/blob/6b7e7ed083882c22b64c01b9318cb57087d45c40/gsadapt_pipeline)

## Acknowledgments

We appreciate the help of Jose Maria Herranz with some bioinformatic analyses and of Augusto Villanueva for providing us with the expanded metadata of the TCGA-LIHC cohort.

