## Supplementary material for "Application of graph models to the identification of transcriptomic oncometabolic pathways in human hepatocellular carcinoma": SupplFigs

**Figure S1. Defining the threshold for the generation of graphs**

**Figure S2. Graph-identified clusters association with iHCC metabolic classes.**

**Figure S3. Selected metabolic signatures according to driver mutation (TP53 or CTNNB1).**

**Figure S4 NME1 signature is upregulated in HCC**

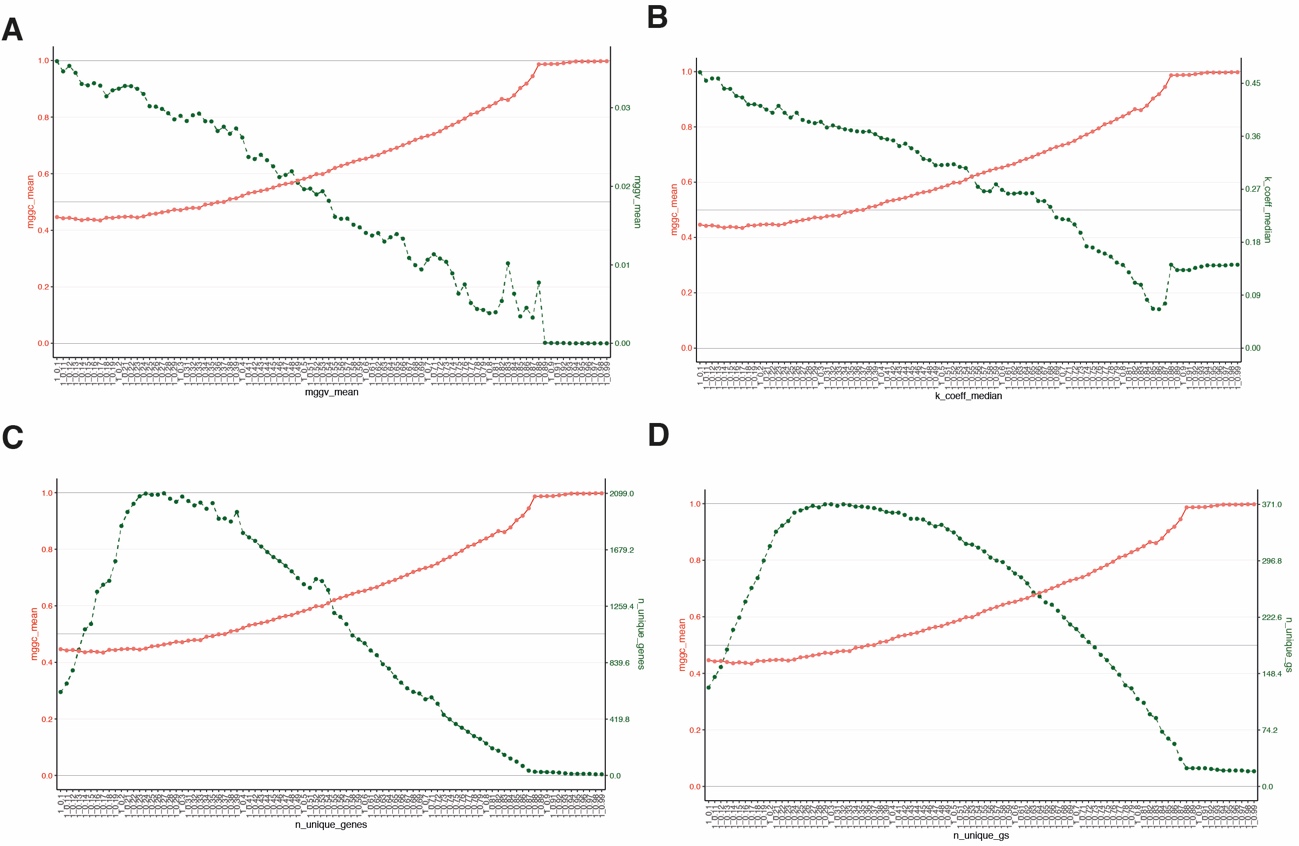

**Figure S1. Defining the threshold for the generation of graphs.** (A) relationship between median gene to gene correlation (MGGC, red dots and line) and median gene to gen variance (MGGV, green dots and line) as the correlation threshold for the matrix of graph generation increases from 0.1 to 1. (B) Relationship between MGGC and K-coefficient for the sequential increase of correlation threshold for graph generation. (C) Same than A but comparing MGGC and number of unique genes included in the signatures. (D) Same than A but comparing MGGC with numer of unique signatures.

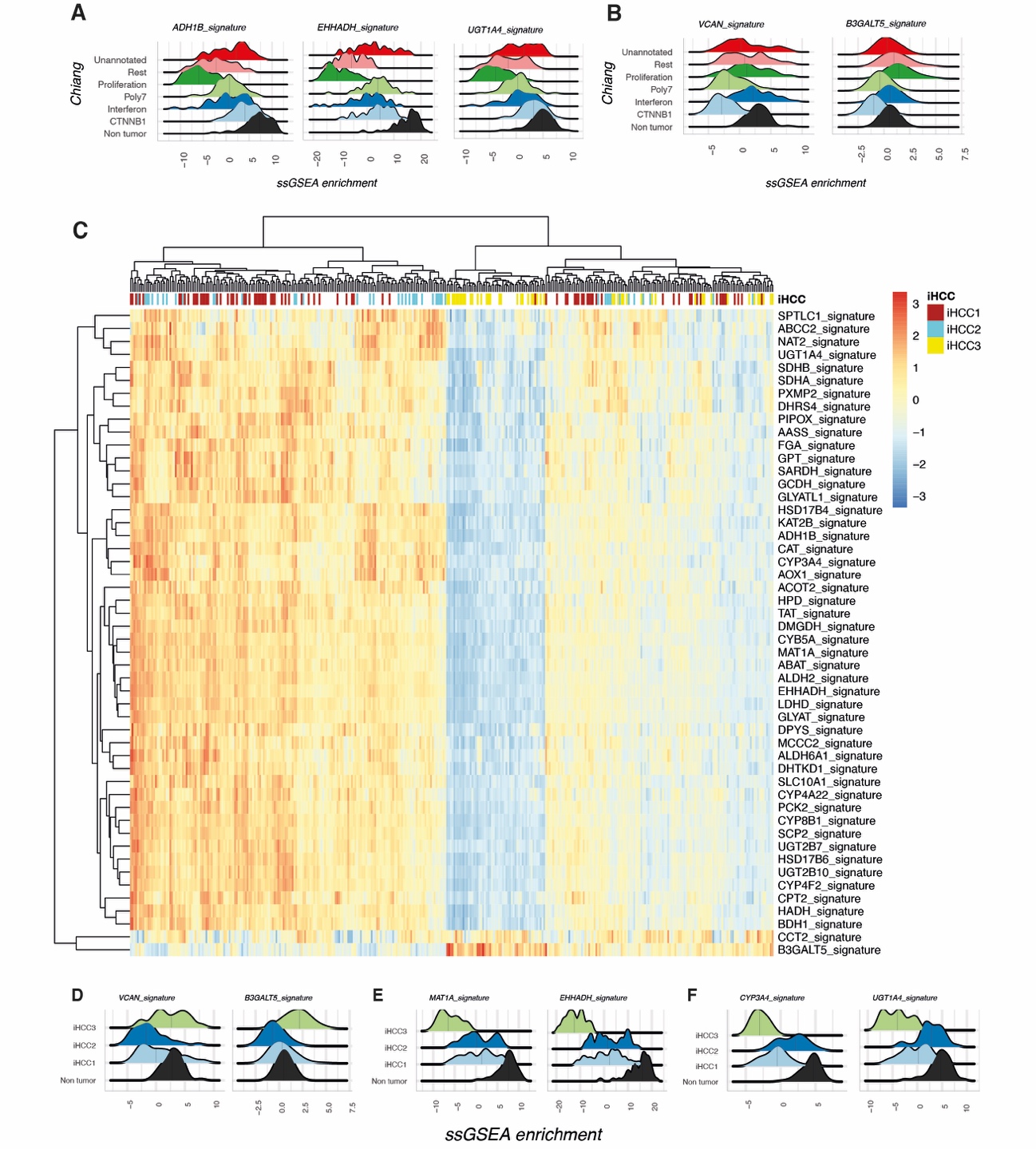

**Figure S2. Graph-identified clusters association with iHCC metabolic classes.**

**(A)** Ridge Plot showing the ssGSEA scores of ADH1B, EHHADH and UGT1A4 signatures, preserved in Chiang classes CTNNB1 and Inteferon and downregulated in Proliferation class. (B) Ridge plot showing the ssGSEA scores of VCAN and B3GALT5 signatures, preserved in Chiang Proliferation class, and downregulated in CTNNB1 class. (C) Heatmap showing the ssGSEA of metabolic clusters identified with the graph modelling, named after the central gene. On top, the identification of patients based on iHCC1, iHCC2 or iHCC3 metabolic classes by Bidkhori et al. (D) Ridge plot showing the ssGSEA scores of VCAN and B3GALT5 signatures, upregulated in iHCC3 subclass patients. (E) Ridge plot showing the ssGSEA scores of MAT1A and EHHADH signatures, downregulated in iHCC3 subclass patients. (F) Ridge plot showing the ssGSEA scores of CYP3A4 and UGT1A4 signatures, specifically upregulated in iHCC2 subclass patients. As in the original work, classes iHCC2 and iHCC1 aggrupated together when compared with iHCC3 subclass.

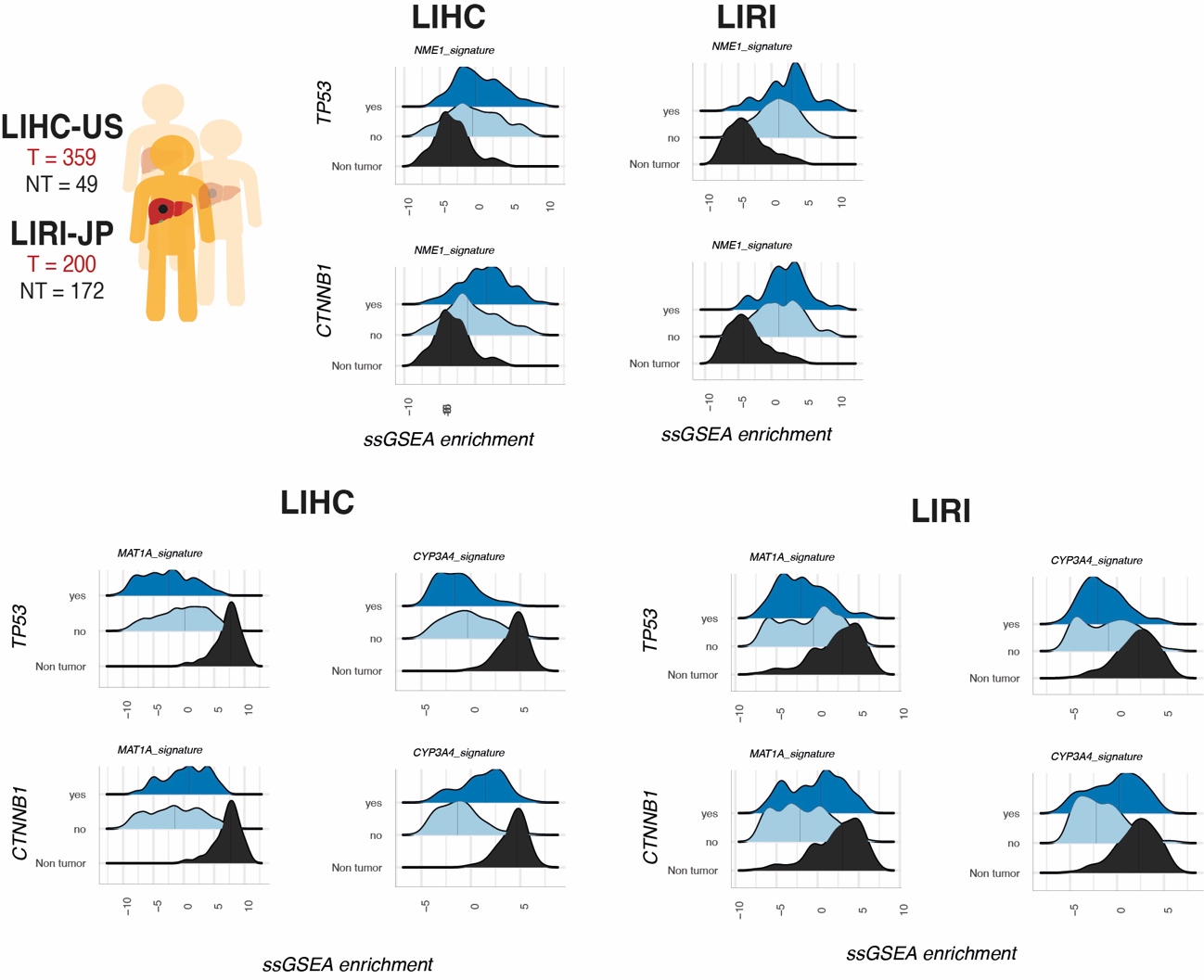

**Figure S3. Selected metabolic signatures according to driver mutation (TP53 or CTNNB1).** ssGSEA scores were compared between TP53 null, CTNNB1 mutated and non tumoral tissue in both LIHC and LIRI cohorts, including NME1 signature (nucleotide metabolism) and MAT1A and CYP3A4 signatures (hepatocyte two-carbon and xenobiotic metabolism).

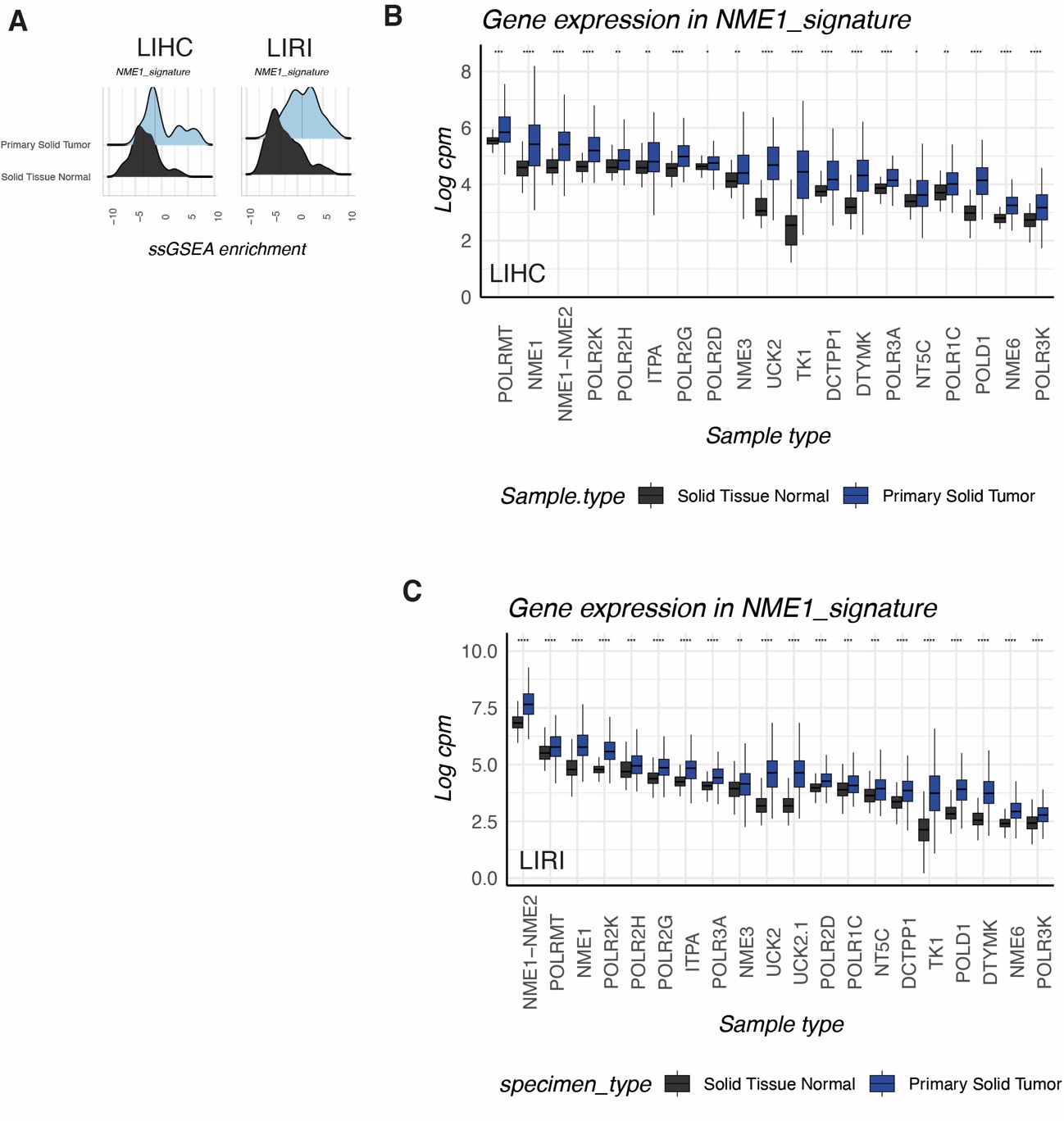

**Figure S4 NME1 signature is upregulated in HCC.** (A) ssGSEA scores of NME signature in HCC cohorts LIRI and LIHC when compared to non tumoral tissue. (B-C) Individual gene expression of components of NME1 signature in LIHC (B) and LIRI-JP (C) cohorts.

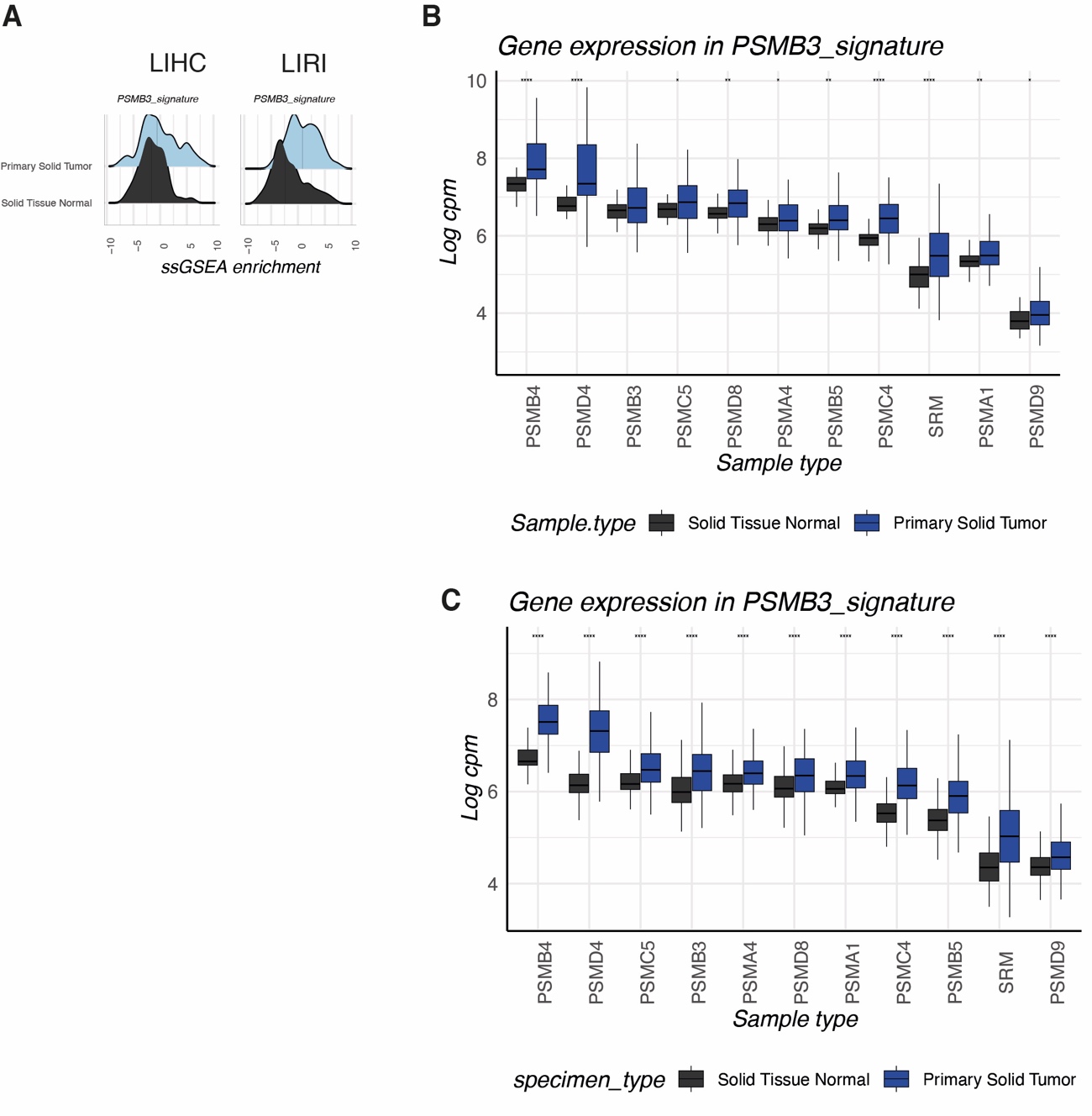

**Figure S5. PSMB3 signature is upregulated in HCC.** (A) ssGSEA scores of PSMB3 signature in HCC cohorts LIRI and LIHC when compared to non tumoral tissue. (B-C) Individual gene expression of components of PSMB3 signature in LIHC (B) and LIRI-JP (C) cohorts.
