## Supplementary material for "Application of graph models to the identification of transcriptomic oncometabolic pathways in human hepatocellular carcinoma": SupplTable1

**Table S1.** **Clinical variables of TCGA-LIHC and ICGC-LIRI-JP cohorts**

| **Clinical variable** | **TCGA-LIHC**  **n=365** | **ICGC-LIRI-JP**  **n=200** | **Test^1^** |
| --- | --- | --- | --- |
| **Tumor type**  Hepatocellular carcinoma  Hepatocellular carcinoma – clear cell type | 361 (98.90%)  4 (1.10%) | 200 (100.00%)  0 (0.00%) | Fisher test  p=0.303 |
| **Gender**  Male  Female | 248 (67.95%)  117 (32.05%) | 150 (75.00%)  50 (25.00%) | Fisher test  p=0.083 |
| **Age**  < 65 years  ≥ 65 years  NA^2^ | 215 (58.90%)  149 (40.82%)  1 (0.30%) | 72 (36.00%)  128 (64.00%)  0 (0.00%) | Fisher test  p<0.001 |
| **Race**  Caucasic – White  Asian  Black or african american  Hispanic or latino  American indian or Alaska native  Not reported | 164 (44.93%)  158 (43.29%)  17 (4.66%)  17 (4.66%)  2 (0.54%)  7 (1.92%) | 0 (0.00%)  200 (100.00%)  0 (0.00%)  0 (0.00%)  0 (0.00%)  0 (0.00%) | Χ^2^ test  p<0.001  **NA out of test* |
| **Hepatitis infection**  HBV  HCV  HBV and HCV  None | 57 (15.62%)  18 (4.93%)  81 (22.19%)  209 (57.26%) | 53 (26.50%)  113 (56.50%)  4 (2.00%)  30 (15.00%) | Χ^2^ test  p<0.001  **NA out of test* |
| **Alcohol consumption**  No or social drinker  Yes  NA | 58 (15.89%)  109 (29.86%)  198 (54.25%) | 108 (54,00%)  83 (41.50%)  9 (4.50%) | Fisher test  p<0.001  **NA out of test* |
| **Edmondson stage**  G1  G2  G3  G4  Undetermined  NA | 53 (14.52%)  173 (47.40%)  122 (33.42%)  12 (3.29%)  0 (0.00%)  5 (1.37%) | 20 (10.00%)  115 (57.50%)  21 (10.50%)  1 (0.5%)  42 (21.00%)  1 (0.5%) | Χ^2^ test  p<0.001  **NA out of test* |
| **TNM stage**  T1  T2  T3  T4  Undetermined  NA | 179 (49.04%)  91 (24.93%)  79 (21.64%)  13 (3.56%)  0 (0.00%)  3 (0.83%) | 19 (9.50%)  112 (56.00%)  20 (10.00%)  1 (0.5%)  47 (23.50%)  1 (0.5%) | Χ^2^ test  p<0.001  **NA out of test* |
| **Hepatic fibrosis**  No fibrosis  Mild fibrosis  Moderate fibrosis  Severe fibrosis  NA | 73 (20.00%)  31 (8.49%)  30 (8.22%)  81 (22.19%)  150 (41.10%) | 10 (5.00%)  27 (13.50%)  40 (20.00%)  123 (61.50%)  0 (0.00%) | Χ^2^ test  p<0.001  **NA out of test* |
| **TP53 mutation**  No  Yes | 256 (70.14%)  109 (29.86%) | 124 (62.00%)  76 (38.00%) | Fisher test  p=0.06 |
| **CTNNB1 mutation**  No  Yes | 270 (73.97%)  95 (26.03%) | 120 (60.00%)  80 (40.00%) | Fisher test  p<0.001 |
| **TERT mutation**  No  Yes | 281 (76.99%)  84 (23.01%) | 175 (87.50%)  25 (12.50%) | Fisher test  p=0.002 |

^1^ Chi Square Test, ^2^  Not Available Data
