## Supplementary material for "Application of graph models to the identification of transcriptomic oncometabolic pathways in human hepatocellular carcinoma": SupplTable2

**Table S2. Characteristics of HCC cell lines from DepMap Portal**

| **ModelID** | **CellLineName** | **DepmapModelType** | **Age** | **AgeCategory** | **Sex** | **PatientRace** | **PrimaryOrMetastasis** | **CCLEName** | **COSMICID** |
| --- | --- | --- | --- | --- | --- | --- | --- | --- | --- |
| ACH-000217 | JHH-6 | HCC | 57 | Adult | Female | asian | Primary | JHH6_LIVER | 1240159 |
| ACH-000221 | SNU-398 | HCC | 42 | Adult | Male | asian | Primary | SNU398_LIVER | 1240217 |
| ACH-000316 | SNU-886 | HCC | 57 | Adult | Male | asian | Primary | SNU886_LIVER | NA |
| ACH-000361 | SK-HEP-1 | HCC | 52 | Adult | Male | caucasian | Metastatic | SKHEP1_LIVER | 909719 |
| ACH-000393 | HLF | HCC | 68 | Adult | Male | asian | Primary | HLF_LIVER | NA |
| ACH-000420 | SNU-449 | HCC | 52 | Adult | Male | asian | Primary | SNU449_LIVER | 909738 |
| ACH-000422 | SNU-475 | HCC | 43 | Adult | Male | asian | Primary | SNU475_LIVER | 909739 |
| ACH-000471 | Li-7 | HCC | 45 | Adult | Male |  | Primary | LI7_LIVER | NA |
| ACH-000475 | huH-1 | HCC | 53 | Adult | Male | asian | Primary | HUH1_LIVER | 1298146 |
| ACH-000476 | JHH-4 | HCC | 51 | Adult | Male | asian | Primary | JHH4_LIVER | 1240158 |
| ACH-000478 | SNU-387 | HCC | 41 | Adult | Female | asian | Primary | SNU387_LIVER | 909736 |
| ACH-000480 | HuH-7 | HCC | 57 | Adult | Male | asian | Primary | HUH7_LIVER | 907071 |
| ACH-000483 | SNU-182 | HCC | 24 | Adult | Male | asian | Primary | SNU182_LIVER | 1240216 |
| ACH-000493 | SNU-423 | HCC | 40 | Adult | Male | asian | Primary | SNU423_LIVER | 909737 |
| ACH-000537 | SNU-761 | HCC | 49 | Adult | Male | asian | Primary | SNU761_LIVER | NA |
| ACH-000577 | JHH-2 | HCC | 57 | Adult | Male | asian | Metastatic | JHH2_LIVER | 1240157 |
| ACH-000620 | JHH-1 | HCC | 50 | Adult | Male | asian | Primary | JHH1_LIVER | 1298151 |
| ACH-000625 | Hep 3B2.1-7 | HCC | 8 | Pediatric | Male | black_or_ african_american | Primary | HEP3B217_LIVER | 1240147 |
| ACH-000686 | SNU-878 | HCC | 54 | Adult | Female | asian | Primary | SNU878_LIVER | NA |
| ACH-000734 | JHH-5 | HCC | 50 | Adult | Male | asian | Primary | JHH5_LIVER | NA |
| ACH-000848 | JHH-7 | HCC | 53 | Adult | Male | asian | Primary | JHH7_LIVER | 1240160 |
| ACH-001089 | HLE | HCC | 68 | Adult | Male | asian | Primary | HLE_LIVER | 907057 |
| ACH-001318 | PLC/PRF/5 | HCC | 24 | Adult | Male |  | Primary | PLCPRF5_LIVER | NA |
| ACH-002523 | SNU-739 | HCC | 52 | Adult | Male |  | Primary | SNU739_LIVER | NA |
