## Supplementary material for "Application of graph models to the identification of transcriptomic oncometabolic pathways in human hepatocellular carcinoma": SupplTable3

**Table S3. Table summary of gene set adaptation in published metabolic signatures**

| **Signature** | **Core** | **Central** | **Original Gene Set** | **MGGC original** | **Size original** | **MGGC adapted** | **Size adapted** |
| --- | --- | --- | --- | --- | --- | --- | --- |
| AASS | AASS, GCDH, ALDH7A1, PIPOX | AASS | REACTOME_LYSINE_CATABOLISM | 0.330 | 12 | 0.549 | 6 |
| ABAT | ABAT, PON3, ALDH6A1, OTC | ABAT | KEGG_ALANINE_ASPARTATE_AND_GLUTAMATE_METABOLISM  WP_GABA_METABOLISM_AKA_GHB  WP_PYRIMIDINE_METABOLISM_AND_RELATED_DISEASES | 0.084 | 45 | 0.476 | 15 |
| ABCC2 | ABCC2, UGT2B4, UGT1A1, UGT1A9, UGT1A6 | ABCC2 | WP_CODEINE_AND_MORPHINE_METABOLISM | 0.319 | 14 | 0.491 | 6 |
| ADCY3 | ADCY3, PDE10A, PDE1A, ENTPD1, PXDN | ADCY3 | Purine metabolism | 0.003 | 157 | 0.322 | 33 |
| ADH1B | ADH1B, ADH6, ALDH2, CAT, CBR1, CYP4F11, MGST1 | ADH1B | Glycolysis – Gluconeogenesis  KEGG_DRUG_METABOLISM_CYTOCHROME_P450  KEGG_RETINOL_METABOLISM  Miscellaneous  Pyruvate metabolism  Serotonin and melatonin biosynthesis  Xenobiotics metabolism | 0.047 | 344 | 0.395 | 72 |
| ALDH2 | ABAT, ACAT1, ACSM2A, ACSM5, ADH1B, ALDH2, ALDH5A1, ALDH6A1, ALDH7A1, ALDH9A1, CAT, CYP4F2, EHHADH, GRHPR, HAGH, LDHD, MAOB, MLYCD, OTC, PCCB, PCK2, RGN | ALDH2 | Ascorbate and aldarate metabolism  Beta-alanine metabolism  KEGG_ARGININE_AND_PROLINE_METABOLISM  KEGG_BETA_ALANINE_METABOLISM  KEGG_BUTANOATE_METABOLISM  KEGG_FATTY_ACID_METABOLISM  KEGG_GLYCOLYSIS_GLUCONEOGENESIS  KEGG_HISTIDINE_METABOLISM  KEGG_PROPANOATE_METABOLISM  KEGG_PYRUVATE_METABOLISM  KEGG_TRYPTOPHAN_METABOLISM  Leukotriene metabolism  Pyruvate metabolism  Tryptophan metabolism | 0.095 | 177 | 0.464 | 58 |
| ALDH6A1 | ALDH6A1, MLYCD, ACADM, DBT, BCKDHB, HIBADH, MCCC2 | ALDH6A1 | Propanoate metabolism  REACTOME_BRANCHED_CHAIN_AMINO_ACID_CATABOLISM | 0.163 | 57 | 0.492 | 18 |
| ALOX5AP | ALOX5AP, ALOX5, TBXAS1, PTGER4, PTGS1 | ALOX5AP | BIOCARTA_EICOSANOID_PATHWAY | 0.256 | 22 | 0.519 | 9 |
| AOX1 | AOX1, MOCS2, SUOX, GPHN, MOCS1, UGT1A4, FMO3 | AOX1 | WP_MOLYBDENUM_COFACTOR_MOCO_BIOSYNTHESIS  WP_NICOTINE_METABOLISM_IN_LIVER_CELLS | 0.418 | 12 | 0.539 | 7 |
| B3GALT5 | B3GALT5, FUT4, B3GNT7, B4GALNT4, GBGT1, GALNT7, B3GNT8 | B3GALT5 | Glycosphingolipid metabolism | 0.015 | 82 | 0.330 | 18 |
| BDH1 | BDH1, ECHDC2, HADH, ACAT1, HMGCL, ECHS1 | BDH1 | Butanoate metabolism  KEGG_BUTANOATE_METABOLISM | 0.149 | 48 | 0.409 | 14 |
| CAT | CAT, HAO1, SCP2, CYP8B1, ABCA6, HSD17B4, AR, AQP9, ALDH9A1 | CAT | HALLMARK_BILE_ACID_METABOLISM | 0.155 | 112 | 0.416 | 34 |
| CHST3 | CHST3, CHST2, B3GNT8, B3GNT7, CHST1 | CHST3 | Keratan sulfate biosynthesis | 0.057 | 23 | 0.407 | 6 |
| COL3A1 | COL3A1, COL5A1, COL1A2, COL6A3, COL6A2, ADAMTS2, COL14A1 | COL3A1 | REACTOME_COLLAGEN_BIOSYNTHESIS_AND_MODIFYING_ENZYMES | 0.094 | 67 | 0.511 | 23 |
| COX5B | COX5B, UQCRQ, UQCR11, NDUFS3, NDUFB7, COX7C, COX6B1, NDUFA2, NDUFA3, COX6A1, COX4I1, NDUFB1, NDUFA13, NDUFA4, COX5A, UQCR10, COX8A, NDUFB8, NDUFB2, COX7B, NDUFA1, NDUFS8, NDUFB10, COX6C, NDUFA8, NDUFA6, NDUFS6, NDUFA12, UQCRC1 | COX5B | Oxidative phosphorylation | 0.399 | 100 | 0.598 | 54 |
| CPT2 | CPT2, SLC25A20, CRAT, ACACB, THRSP, PRKAG2 | CPT2 | Carnitine shuttle (mitochondrial)  REACTOME_CARNITINE_METABOLISM | 0.091 | 20 | 0.497 | 7 |
| CYB5A | ALDH9A1, CTH, CYB5A, CYP8B1, GYS2, HSD11B1, MTHFD1, PC, PCCB, POR, RBP4, SDC1, SOD1, SRD5A2 | CYB5A | HALLMARK_GLYCOLYSIS  REACTOME_METABOLISM_OF_VITAMINS_AND_COFACTORS  WP_CLASSICAL_PATHWAY_OF_STEROIDOGENESIS_WITH_GLUCOCORTICOID_AND_MINERALOCORTICOID_METABOLISM | 0.037 | 396 | 0.329 | 107 |
| CYP3A4 | CYP1A2, CYP2C9, CYP3A4, FMO3, UGT1A4 | CYP3A4 | BIOCARTA_NUCLEARRS_PATHWAY  REACTOME_BIOSYNTHESIS_OF_MARESIN_LIKE_SPMS  REACTOME_BIOSYNTHESIS_OF_MARESINS  REACTOME_BIOSYNTHESIS_OF_SPECIALIZED_PRORESOLVING_MEDIATORS_SPMS  WP_AFLATOXIN_B1_METABOLISM  WP_TAMOXIFEN_METABOLISM | 0.156 | 68 | 0.471 | 15 |
| CYP4A22 | CYP4A22, CYP4A11, CYP2C8, CYP4F2 | CYP4A22 | WP_EICOSANOID_METABOLISM_VIA_CYTOCHROME_P450_MONOOXYGENASES_PATHWAY | 0.476 | 9 | 0.516 | 7 |
| CYP4F2 | ACAA2, ACOX1, BAAT, CBR1, CYP2A6, CYP2C8, CYP4A11, CYP4A22, CYP4F2, CYP4F3, DHRS4, GSTK1, GSTZ1, HSD17B10, MGST1, SLC27A2 | CYP4F2 | Arachidonic acid metabolism  Drug metabolism  Eicosanoid metabolism  KEGG_ARACHIDONIC_ACID_METABOLISM | 0.066 | 183 | 0.413 | 56 |
| CYP8B1 | ABCB11, ABCB4, ACOX2, CYP2J2, CYP4A11, CYP4A22, CYP4F2, CYP8B1, NR1I2, PON1, SCL10A1, SCP2, SLC27A5 | CYP8B1 | BIOCARTA_NUCLEARRS_PATHWAY  KEGG_PRIMARY_BILE_ACID_BIOSYNTHESIS  REACTOME_ARACHIDONIC_ACID_METABOLISM  REACTOME_BILE_ACID_AND_BILE_SALT_METABOLISM  WP_NUCLEAR_RECEPTORS_IN_LIPID_METABOLISM_AND_TOXICITY | 0.082 | 128 | 0.476 | 45 |
| DCN | DCN, BGN, VCAN, DSE | DCN | REACTOME_DERMATAN_SULFATE_BIOSYNTHESIS | 0.117 | 11 | 0.497 | 6 |
| DHCR7 | DHCR7, EBP, LSS, NSDHL, MSMO1 | DHCR7 | Cholesterol biosynthesis 1 (Bloch pathway)  KEGG_STEROID_BIOSYNTHESIS  WP_CHOLESTEROL_BIOSYNTHESIS_WITH_SKELETAL_DYSPLASIAS | 0.342 | 22 | 0.463 | 11 |
| DHRS4 | DHRS4, GSTK1, ECHS1, CBR1, MGST1, CYP4F11, HSD17B10, EPHX1 | DHRS4 | Arachidonic acid metabolism | 0.110 | 111 | 0.369 | 24 |
| DHTKD1 | AASS, ALDH9A1, BCKDHB,  DBT, DHTKD1, GOT2, SETD7 | DHTKD1 | Lysine metabolism  REACTOME_GLYOXYLATE_METABOLISM_AND_GLYCINE_DEGRADATION | 0.052 | 76 | 0.407 | 18 |
| DLD | DLD, DLST, SUCLG2P2, ACO2, SDHC | DLD | KEGG_CITRATE_CYCLE_TCA_CYCLE  KEGG_GLYCOLYSIS_GLUCONEOGENESIS | 0.076 | 86 | 0.307 | 25 |
| DMGDH | ALDH7A1, BHMT, CTH, DAO, DMGDH, MAOB, SARDH, SHMT1 | DMGDH | KEGG_GLYCINE_SERINE_AND_THREONINE_METABOLISM  REACTOME_CHOLINE_CATABOLISM | 0.319 | 33 | 0.539 | 14 |
| DPYS | DPYS, XDH, UPB1, AGXT2 | DPYS | REACTOME_NUCLEOTIDE_CATABOLISM | 0.029 | 35 | 0.395 | 9 |
| EBP | EBP, NSDHL, TM7SF2, HSD17B7 | EBP | Cholesterol biosynthesis 3 (Kandustch-Russell pathway)  KEGG_STEROID_BIOSYNTHESIS | 0.229 | 12 | 0.535 | 4 |
| EHHADH | EHHADH, ACAA1, ACOX1, SLC27A5, CYP8B1, ALDH2, HSD17B4, CDO1, SCP2, ACAA2, CPT2, ETFDH, ETFA, ACOX2, CYP4F2, SLC27A2, HAO1, CAT, ABAT, ALDH6A1, SUCLG2, ALDH9A1, ECI2, MLYCD, ACBD4, HADH, HSD11B1, IVD, HIBADH, OTC, CYP4A22, CYP4A11 | EHHADH | Beta oxidation of even-chain fatty acids (peroxisomal)  Beta oxidation of phytanic acid (peroxisomal)  Bile acid biosynthesis  Drug metabolism  Fatty acid oxidation  HALLMARK_FATTY_ACID_METABOLISM  KEGG_FATTY_ACID_METABOLISM  KEGG_PEROXISOME  KEGG_PROPANOATE_METABOLISM  Omega-3 fatty acid metabolism  REACTOME_FATTY_ACID_METABOLISM  REACTOME_PEROXISOMAL_LIPID_METABOLISM  Steroid metabolism  Valine; leucine; and isoleucine metabolism  WP_AMINO_ACID_METABOLISM  WP_EICOSANOID_METABOLISM_VIA_LIPOOXYGENASES_LOX | 0.106 | 562 | 0.444 | 189 |
| EP300 | CREBBP, EP300, GABPA, MEF2A , SIRT1 | EP300 | REACTOME_REGULATION_OF_LIPID_METABOLISM_BY_PPARALPHA  WP_ENERGY_METABOLISM | 0.037 | 156 | 0.340 | 40 |
| FDPS | FDPS, MVD, HSD17B7, EBP | FDPS | REACTOME_CHOLESTEROL_BIOSYNTHESIS | 0.388 | 27 | 0.514 | 8 |
| FGA | FGA, FGB, FGG, MAT1A, SERPINA1 | FGA | Protein assembly  WP_FOLATE_METABOLISM | 0.105 | 77 | 0.416 | 24 |
| GCDH | GCDH, ACAT1, HAAO, ALDH7A1 | GCDH | KEGG_TRYPTOPHAN_METABOLISM | 0.187 | 40 | 0.447 | 10 |
| GLYAT | ALDH1B, ALDH2, CAT, DMGDH, GRHPR, GLYAT, GLYATL1, HAO1, HGD, HPD, TAT, UGT2B10 | GLYAT | Glycine; serine and threonine metabolism  Phenylalanine; tyrosine and tryptophan biosynthesis | 0.033 | 272 | 0.397 | 63 |
| GLYATL1 | GLYATL1, PIPOX, DAO, SARDH | GLYATL1 | Glycine; serine and threonine metabolism | 0.098 | 61 | 0.468 | 16 |
| GMPPA | GMPPA, ALG3, DPM3, NANS, DPM2 | GMPPA | REACTOME_BIOSYNTHESIS_OF_THE_N_GLYCAN_PRECURSOR_DOLICHOL_LIPID_LINKED_OLIGOSACCHARIDE_LLO_AND_TRANSFER_TO_A_NASCENT_PROTEIN | 0.036 | 78 | 0.267 | 18 |
| GNB4 | GNB4, IQGAP1, GNG2, ADCY3, CACNA1C | GNB4 | REACTOME_INTEGRATION_OF_ENERGY_METABOLISM | 0.017 | 108 | 0.365 | 23 |
| GPT | GPT, GOT2, AGXT, ASS1, CPS1, GPT2, ASL | GPT | KEGG_ALANINE_ASPARTATE_AND_GLUTAMATE_METABOLISM  WP_ALANINE_AND_ASPARTATE_METABOLISM | 0.12 | 34 | 0.521 | 11 |
| GPX4 | GPX1, GPX3, GPX4, GSS, PRDX5, PRDX6 | GPX4 | Glutathione metabolism  WP_GLUTATHIONE_METABOLISM | 0.051 | 41 | 0.410 | 8 |
| HADH | HADH, ECHS1, ACAA2, EHHADH, ACAA1, HSD17B4, ACAT1, GSTK1, GSTZ1, MGST1, ECHDC2, ECHDC3 | HADH | Fatty acid oxidation  Omega-6 fatty acid metabolism  Tryptophan metabolism  Vitamin E metabolism  WP_FATTY_ACID_BIOSYNTHESIS | 0.237 |  | 0.436 | 50 |
| HMGCL | HMGCL, GCDH, ACAA1, ECHS1, ECI2 | HMGCL | HALLMARK_FATTY_ACID_METABOLISM | 0.129 | 158 | 0.315 | 39 |
| HPD | HGD, HPD, MAOB, QDPR | HPD | KEGG_TYROSINE_METABOLISM  REACTOME_PHENYLALANINE_AND_TYROSINE_METABOLISM | 0.124 | 47 | 0.512 | 13 |
| HSD17B4 | ACAA1, ACSL5, AMACR, HSD17B4, MLYCD | HSD17B4 | Beta oxidation of phytanic acid (peroxisomal)  REACTOME_PEROXISOMAL_LIPID_METABOLISM | 0.308 | 42 | 0.469 | 15 |
| HSD17B6 | HSD17B6, SRD5A2, PIPOX, NR1I2, CES2, UGT2B10, UGT2B7, RBP4 | HSD17B6 | Androgen metabolism  HALLMARK_BILE_ACID_METABOLISM  Retinol metabolism  Steroid metabolism | 0.109 |  | 0.376 | 68 |
| IDI1 | IDI1, HMGCR, HMGCS1, FDPS, ACAT2 | IDI1 | Cholesterol biosynthesis 1 (Bloch pathway)  Cholesterol metabolism  REACTOME_CHOLESTEROL_BIOSYNTHESIS  WP_MEVALONATE_ARM_OF_CHOLESTEROL_BIOSYNTHESIS_PATHWAY | 0.157 | 45 | 0.525 | 22 |
| ITPKB | ITPKB, INPP4B, INPP5D, PLCB2, PLCB4 | ITPKB | REACTOME_INOSITOL_PHOSPHATE_METABOLISM | 0.038 | 48 | 0.362 | 10 |
| KAT2B | KAT2B, CAT, ALDH6A1, ALAD | KAT2B | HALLMARK_HEME_METABOLISM | 0.025 | 200 | 0.242 | 52 |
| LDHD | LDHD, HAO1, ALDH2, SDHD, ALDH9A1, GRHPR | LDHD | Tricarboxylic acid cycle and glyoxylate-dicarboxylate metabolism | 0.098 | 49 | 0.479 | 17 |
| LUM | LUM, PRELP, DCN, BGN, FMOD | LUM | REACTOME_DISEASES_ASSOCIATED_WITH_GLYCOSAMINOGLYCAN_METABOLISM | 0.069 | 41 | 0.698 | 10 |
| MAT1A | MAT1A, TAT, PON3, CTH, FMO4, CDO1, BHMT, SUOX, SAA4 | MAT1A | Cysteine and methionine metabolism  Metabolism of other amino acids  REACTOME_METABOLISM_OF_INGESTED_SEMET_SEC_MESEC_INTO_H2SE  WP_CYSTEINE_AND_METHIONINE_CATABOLISM  WP_METHIONINE_METABOLISM_LEADING_TO_SULFUR_AMINO_ACIDS_AND_RELATED_DISORDERS  WP_VITAMIN_B12_METABOLISM | 0.103 | 126 | 0.430 | 40 |
| MCCC2 | MCCC2, PCCB, PC, PCCA | MCCC2 | REACTOME_DEFECTS_IN_BIOTIN_BTN_METABOLISM | 0.388 | 8 | 0.456 | 6 |
| NAT2 | NAT2, CES1, UGT1A4, UGT1A3, CYP3A4 | NAT2 | KEGG_DRUG_METABOLISM_OTHER_ENZYMES | 0.107 | 51 | 0.452 | 13 |
| NME1 | NME1, POLR2H, POLR2J, POLR2I, ITPA, NT5C, POLR2L | NME1 | Nucleotide metabolism | 0.009 | 156 | 0.301 | 36 |
| PCK2 | PCK2, SLC27A5, EHHADH, CYP8B1, SCP2, CYP4A22, ACOX2 | PCK2 | KEGG_PPAR_SIGNALING_PATHWAY | 0.085 | 69 | 0.460 | 28 |
| PIK3C2A | PIK3C2A, PIKFYVE, SYNJ1, PIK3CA, PIK3R4 | PIK3C2A | KEGG_INOSITOL_PHOSPHATE_METABOLISM  REACTOME_PI_METABOLISM  WP_PHOSPHOINOSITIDES_METABOLISM | 0.068 | 77 | 0.348 | 30 |
| PIPOX | PIPOX, SARDH, GLYCTK, AGXT | PIPOX | KEGG_GLYCINE_SERINE_AND_THREONINE_METABOLISM | 0.251 | 31 | 0.490 | 8 |
| PSMB3 | PSMB3, PSMD13, PSMC3, PSMA7, PSMC5, PSMB4, PSMC4, PSMD8 | PSMB3 | REACTOME_METABOLISM_OF_POLYAMINES | 0.205 | 59 | 0.507 | 20 |
| PXMP2 | PXMP2, ECI2, PEX11G, HMGCL, GSTK1, DHRS4 | PXMP2 | KEGG_PEROXISOME | 0.193 | 78 | 0.442 | 19 |
| RPL37A | RPL37A, RPL32, RPL24, RPS14, RPL31, RPS9, RPL37, RPL29, RPL14, RPL36, RPS13, RPL27, RPL18, RPL38, RPS11, RPL19, RPL27A, RPS16, RPLP2, RPL35A, UBA52, RPL18A, RPL23, RPS8, RPL35, RPS17, RPS21, RPL28, RPS28, RPL6, RPL13A, RPL23A, RPS27A, RPL12, RPL13, RPL34, RPS23, FAU, RPS10, RPL7A, RPS7, RPS15A, RPS24, RPL11, RPL36A, RPLP1, RPL39, RPS5, RPS3 | RPL37A | REACTOME_METABOLISM_OF_AMINO_ACIDS_AND_DERIVATIVES | 0.051 | 374 | 0.513 | 115 |
| SARDH | ASL, ASS1, CPS1, DMGDH, GOT1, HNMT, SARDH | SARDH | Sulfur metabolism  WP_UREA_CYCLE_AND_METABOLISM_OF_AMINO_GROUPS | 0.046 | 23 | 0.478 | 7 |
| SCP2 | SCP2, SAR1B, CYP8B1, SC5D, SLC27A5, HSD17B4, ABCB11, ACOX2, SLC10A1, SLC27A2, INSIG1, PPARA | SCP2 | REACTOME_ALPHA_LINOLENIC_OMEGA3_AND_LINOLEIC_OMEGA6_ACID_METABOLISM  REACTOME_METABOLISM_OF_STEROIDS | 0.070 | 165 | 0.443 | 43 |
| SDHA | ASS1, CPS1, DMGDH, GOT1, HNMT, SARDH | SDHA | BIOCARTA_KREB_PATHWAY  REACTOME_PYRUVATE_METABOLISM_AND_CITRIC_ACID_TCA_CYCLE | 0.045 | 33 | 0.478 | 8 |
| SDHB | SDHB, SDHA, UQCRC1, SDHD, SLC25A4 | SDHB | BIOCARTA_ETC_PATHWAY | 0.303 | 10 | 0.405 | 7 |
| SLC10A1 | SLC10A1, SLC27A5, ABCB11, SLC27A2 | SLC10A1 | Bile acid recycling | 0.187 | 16 | 0.443 | 7 |
| SPTLC1 | SPTLC1, TMPPE, ST6GALNAC3, ACER2 | SPTLC1 | Sphingolipid metabolism | 0.016 | 141 | 0.092 | 29 |
| TAT | TAT, CTH, MAT1A, HPD, PAH, MAOB, GOT2, ALDH7A1 | TAT | Alanine; aspartate and glutamate metabolism  KEGG_CYSTEINE_AND_METHIONINE_METABOLISM  KEGG_PHENYLALANINE_METABOLISM  WP_AMINO_ACID_METABOLISM | 0.061 | 159 | 0.466 | 37 |
| THBS2 | THBS2, LUM, ADAMTS2, DCN, PRELP, VCAN, ADAMTS12, NOTCH3, FMOD, BGN | THBS2 | REACTOME_DISEASES_OF_METABOLISM | 0.015 | 250 | 0.321 | 60 |
| UGT1A4 | UGT1A4, UGT1A3, UGT2B10, UGT2B7, MGST1, CYP1A2, CYP1A1, UGT2B4, GBE1, ALAS1, ABCG2, NAT2 | UGT1A4 | KEGG_ASCORBATE_AND_ALDARATE_METABOLISM  KEGG_METABOLISM_OF_XENOBIOTICS_BY_CYTOCHROME_P450  KEGG_PORPHYRIN_AND_CHLOROPHYLL_METABOLISM  KEGG_STARCH_AND_SUCROSE_METABOLISM  KEGG_STEROID_HORMONE_BIOSYNTHESIS  REACTOME_METABOLISM_OF_PORPHYRINS  WP_ARYLAMINE_METABOLISM | 0.129 | 171 | 0.420 | 42 |
| UGT2B10 | UGT2B10, UGT2B7, CYP2C8, ADH1C, CYP3A4, CYP4A22, RDH16, CYP2A6, ADHFE1, HSD11B1, CYP2J2, SULT2A1 | UGT2B10 | KEGG_DRUG_METABOLISM_CYTOCHROME_P450  KEGG_METABOLISM_OF_XENOBIOTICS_BY_CYTOCHROME_P450  KEGG_RETINOL_METABOLISM  Linoleate metabolism  Xenobiotics metabolism | 0.124 | 162 | 0.376 | 45 |
| UGT2B7 | UGT2B7, GYS2, UGT2B10, GBA3, HSD17B6, CYP3A4, HSD11B1, SRD5A2, UGT2B15, SLCO1B1, CYP2C9, CYP2C8 | UGT2B7 | KEGG_STARCH_AND_SUCROSE_METABOLISM  KEGG_STEROID_HORMONE_BIOSYNTHESIS  WP_CODEINE_AND_MORPHINE_METABOLISM  WP_TAMOXIFEN_METABOLISM | 0.147 | 104 | 0.441 | 31 |
| VCAN | VCAN, FMOD, LUM, XYLT1, SLC9A1, BGN, CHST3 | VCAN | REACTOME_CHONDROITIN_SULFATE_BIOSYNTHESIS  REACTOME_CHONDROITIN_SULFATE_DERMATAN_SULFATE_METABOLISM  REACTOME_METABOLISM_OF_CARBOHYDRATES | 0.017 | 246 | 0.371 | 53 |
